# The LV-LA Health Score: A Novel Marker of Integrated Myocardial Structure and Function

**DOI:** 10.64898/2026.06.08.26353379

**Authors:** Frank Estrella, Karen Chiswell, Jie-Lena Sun, Shelly Duckworth, Ramachandran S. Vasan, Brenda Pattinson, Alicia Provencher, Suzanne E. Judd, Raghava Velagaleti, Pamela Douglas, Gerald Bloomfield, Elsayed Soliman, Yii-Der Ida Chen

**Affiliations:** Duke University Hospital, Durham, NC; Duke Clinical Research Institute, Durham, NC; University of Texas Health, San Antonio, TX; University of Alabama at Birmingham, Birmingham, AL; Signature Healthcare, Brockton, MA; Wake Forest University School of Medicine, Winston-Salem, NC; The Lundquist Institute for Biomedical Innovation at Harbor-UCLA Medical Center, Torrance, CA

## Abstract

**Background:** Myocardial remodeling precedes symptomatic heart failure, which is important to detect early. We assessed feasibility and clinical correlates of a novel integrated assessment of myocardial remodeling in a large rural cohort in the Southeastern United States.

**Methods:** Echoes were obtained with AI assistance (Caption guidance) in 3100 adults in the NHLBI-funded RURAL cohort study. Of those, 1895 had quantifiable global longitudinal strain (GLS), left ventricular mass (LVM), and left atrial volume (LAV). LV-LA Health was based on a simple count of sex-specific abnormalities (0-3), indexed to body surface area (BSA) or height (Table 1). Relationships with demographics and risk factors were compared with Spearman correlation and Mantel-Haenszel tests, with moderate and severe results combined.

**Results:** Median (IQR) age was 49 (40-58). Impaired LV-LA Health is common even in a low PREVENT cardiovascular (CV) risk population (median 10-year risk 3.3%; 25th, 75th 1.2,7.2) with preserved ejection fraction (EF; 60%; 57,62). The prevalence of abnormalities differed greatly by indexing method: 18.2% with BSA (15.1% mild; 3.1% mod/severe) vs 51% with height (38.3% mild; 12.7% mod/severe) (Figure 1). LV-LA impairment increased with age, PREVENT CV risk score and cardiovascular risk factors (hypertension, diabetes, dyslipidemia, obesity); all p<0.001. Impairment was more common in Black vs White people (p<0.001) and differed by sex only with height indexation.

**Conclusions:** A novel LV-LA health composite of routinely acquired echocardiographic measures identifies substantial subclinical cardiac remodeling in a middle-aged rural community cohort, not detected by PREVENT score or ejection fraction. This is the first application of this framework in a large, unselected community sample. Indexation method affects prevalence, with BSA likely underestimating risk in adiposity-enriched populations. Findings suggest a high rural burden and longitudinal evaluation with future CV events is ongoing.

## Introduction

Heart failure (HF) remains a major global health burden affecting more than 64 million people with projections of prevalence and cost continuing to increase through 2030 and beyond (1,2). Detecting myocardial dysfunction during the asymptomatic phase (Stage A/B HF) has become central to preventive cardiology. The heart will experience progressive myocardial remodeling years before symptoms develop, yet our conventional measures of heart failure such as left ventricular ejection fraction % (LVEF%), serum biomarkers (BNP, troponin), identify dysfunction only after significant damage has occurred which precludes opportunities for early interventions (3–5). Cardiovascular disease (CVD) disproportionately affects rural communities in the southern United States, where limited access and greater social stressors contribute to increased prevalence and excess mortality with rural adults having a 19% higher incidence of HF than urban populations (6,7).

Advances in echocardiography now enable a more nuanced assessment of myocardial health. Left ventricular mass index (LVMi) and left atrial volume index (LAVi) capture complementary domains of remodeling and reflect adaptive responses to chronic hemodynamic stress and cumulative diastolic burden, respectively (8,9). Global longitudinal strain (GLS), derived from speckle-tracking echocardiography, is a sensitive marker of early contractile dysfunction, with modest reductions (absolute GLS <15–18%) associated with increased cardiovascular risk, even with normal LVEF (10). Abnormalities across multiple domains provide incremental prognostic value beyond individual measures (11,12), and improvement in these parameters have been associated with more favorable outcomes in interventional settings such as valve replacement (13). Although body surface area (BSA) indexation remains the guideline standard, it may underestimate remodeling in individuals with obesity, whereas height-based indexation may better capture these changes (14,15).

In this study, we apply a LV–LA Health phenotype using LVMi and LAVi as primary structural measures of myocardial remodeling. We aim to (1) determine the feasibility of constructing this phenotype in a large rural cohort, (2) describe its cross-sectional distribution across alternative indexation strategies, and (3) evaluate associations with cardiometabolic risk factors and demographic characteristics. We further examine whether differences in body size indexation meaningfully influence phenotype classification and risk associations.

## Methods

### Study Design and Population

This study is a cross-sectional analysis of participants enrolled in the echocardiographic sub-study of the Risk Underlying Rural Areas Longitudinal (RURAL) study (RURAL ECHO), an NHLBI-funded, community-based cohort of adults residing in rural counties across Kentucky, Alabama, Mississippi, and Louisiana. The RURAL study was designed to evaluate cardiovascular risk in a multiethnic population sampled from counties stratified by mortality risk and ecologically paired by poverty level, race/ethnic composition, and population size. ** Statement about IRB here**

### Inclusion and exclusion criteria

Participants were eligible for inclusion if they were ≥18 years of age, enrolled in the parent RURAL study, and had quantifiable left ventricular (LV) mass, left atrial (LA) volume, and anthropometric data (height and weight) required for indexation. Exclusion criteria included moderate or severe valvular heart disease, congenital heart disease, or inadequate echocardiographic image quality precluding reliable measurement.

Of 3,833 participants with an attempted echocardiographic examination, 3,513 had complete LV mass, LA volume, and anthropometric data and comprised the primary analytic cohort. The proportion of participants in whom a complete LV–LA Health phenotype could be constructed was 92% (Table 0).

### Echocardiographic Measurements

Transthoracic echocardiography was performed using standardized acquisition protocols with AI-assisted image guidance (Caption Guidance) and real-time quality assurance. Measurements followed American Society of Echocardiography (ASE) and European Association of Cardiovascular Imaging (EACVI) recommendations for chamber quantification and strain analysis.

Left ventricular mass (LVM) was calculated from two-dimensional echocardiographic measurements using ASE-recommended linear dimension–based methods (area–length approach) and expressed in grams. LAV was obtained using the biplane area-length method via 2D echo when available, followed by single plane and linear measurements in a hierarchical fashion only if needed and expressed in milliliters. LVM and LAV were indexed to body surface area (BSA) and height²·⁷ for the inferential analyses.

Additional indexation strategies were evaluated in descriptive analyses only. These included LVM indexed to height²·⁷ combined with LAV indexed to height²·⁷, height², and height¹·⁷², as well as mixed indexation approaches including LVM indexed to BSA with LAV indexed to height²·⁷, height², and height¹·⁷². These descriptive analyses were used to assess differences in LV–LA Health phenotype distribution and quantify reclassification relative to the BSA-indexed reference.

Abnormal cut points for BSA-indexed measures were derived from guideline-recommended thresholds, while height-based thresholds were based on values in prior literature (15).

Global longitudinal strain (GLS) was derived from three apical views using vendor-independent speckle-tracking software and reported as an absolute value. Due to substantial missingness, GLS was not included in the primary LV–LA Health phenotype and was instead evaluated in supplemental descriptive analyses.

### LV–LA Health Phenotype Definition

The LV–LA Health phenotype was defined as a composite measure of structural cardiac remodeling using indexed LVM and LAV. For each participant, binary indicators were created for abnormal LVM and abnormal LAV based on prespecified cut points (Cut point table).

The LV–LA Health phenotype was defined ordinally based on the number of abnormal indices, with participants classified as having preserved health when no abnormalities were present, mildly impaired health when one abnormal index was identified (either LVMi or LAVi), and moderately to severely impaired health when both indices were abnormal. The primary phenotype definition used BSA-indexed LVM and LAV since this is the most validated indexation method. A parallel phenotype was constructed using height²·⁷ indexation for both

LVM and LAV for comparative analyses. Additional height-based indexation strategies were evaluated descriptively.

Supplemental analyses incorporating GLS was performed to further stratify impairment severity; however, these were not included in the primary phenotype due to incomplete data availability.

### Clinical Covariates

All clinical data was obtained through the RURAL cohort. Demographic variables included age, sex, race/ethnicity, and Hispanic/Latino ethnicity. County-level cardiovascular risk was categorized based on residence in high-risk versus low-risk counties, where high-risk counties were defined in the RURAL study as those with approximately 70% higher-than-average heart, lung, blood, and sleep-related mortality, and low-risk counties as those with approximately 20% lower-than-average mortality. Cardiometabolic risk factors included hypertension (AHA/ACC 2017 definition, blood pressure ≥130/80 mmHg or use of antihypertensive medications), dyslipidemia (total cholesterol >200 mg/dL, LDL >130 mg/dL, triglycerides >150 mg/dL, or HDL <40-50 mg/dL or presence cholesterol lowering medication), diabetes mellitus (hemoglobin A1c >/= 6.5% or presence of anti-hyperglycemic medication), obesity, smoking status, family history and overall cardiovascular risk burden (PREVENT 10-year total cardiovascular disease risk score (%). Echocardiographic measures were analyzed alongside demographic and clinical variables (Table)

## Statistical Analysis

Baseline demographic, clinical, and echocardiographic variables were obtained from the RURAL study baseline examination and corresponding echocardiographic assessment. Characteristics were summarized for the overall analytic cohort and stratified by LV–LA Health phenotype category. Continuous variables were described using means with standard deviations or medians with interquartile ranges, and categorical variables were summarized as counts and percentages. Feasibility of the LV–LA Health phenotype was defined as the proportion of participants with complete left ventricular mass (LVM), left atrial volume (LAV), and anthropometric data required for indexation. Phenotype distributions were described overall and across indexation strategies.

### Descriptive Comparisons

Baseline demographic, clinical, and echocardiographic characteristics were summarized by LV–LA Health phenotype category for both BSA and height²·⁷ indexation. Group differences were assessed using Spearman rank correlation tests for continuous variables and Mantel–Haenszel chi-square tests for categorical variables

### Association Analyses

To evaluate associations between cardiometabolic risk factors and LV–LA Health phenotype, multinomial logistic regression models were fitted with phenotype category (preserved, mildly impaired, moderately/severely impaired) as the dependent variable and preserved LV–LA health as the reference group. Prespecified exposures included age, hypertension, diabetes mellitus, dyslipidemia, smoking status, body mass index, sex, race/ethnicity, family history of cardiovascular disease, and county-level cardiovascular risk. Three sequential modeling approaches were used. First, univariable multinomial logistic regression models were fitted for each exposure separately to estimate unadjusted associations. Second, models adjusted for age were constructed to account for the strong confounding effect of age on myocardial remodeling. Third, fully adjusted multivariable models were fitted including all prespecified exposures to estimate independent associations with LV–LA Health phenotype. Continuous variables were modeled as linear terms, including age per 10-year increase and body mass index per 1 kg/m² increase. Model results are reported as odds ratios (ORs) with 95% confidence intervals (CIs).

Primary analyses were performed using body surface area (BSA)-indexed LVM and LAV. A secondary inferential analysis was conducted using height²·⁷ indexation for both measures to directly compare results with the BSA-based approach. Additional indexation strategies, including LAV indexed to height² and height¹·⁷² and mixed combinations of BSA and height-based scaling, were evaluated descriptively to assess differences in LV–LA Health phenotype distribution.

### Indexation Comparisons and Reclassification

To further evaluate the impact of indexation strategy, reclassification analyses were performed comparing alternative indexation approaches with the BSA-based reference. Reclassification was quantified as the proportion of participants reassigned to higher or lower phenotype severity categories, and net reclassification was calculated as the difference between these proportions.

### Biomarker Analyses Missing Data

Missing covariate data were handled using single imputation, with median values used for continuous variables and mode values for categorical variables. All analyses were conducted using two-sided testing with a significance level of α = 0.05.

## Results

### Analytic cohort and feasibility

Among 3,833 RURAL ECHO participants with an attempted echocardiographic examination, 101 were excluded due to missing LA volume and 219 due to missing LV mass. No additional participants were excluded for missing BSA or height, yielding a primary analytic cohort of 3,513 participants with complete LVM, LAV, and anthropometric data. The LV–LA Health phenotype was feasible in 92% of participants **(Table 0)**

In the overall cohort, the median age (IQR) was 49 (40–57) years, 68% were female, and 58% identified as White with 41% identifying as Black. Obesity was present in 59% of participants, with a median BMI (IQR) of 32 kg/m² (27–38). Cardiometabolic risk factors were common, including hypertension (69%) and dyslipidemia (47%). Mean left ventricular ejection fraction was 59%, and median (IQR) PREVENT score was 3.3% (1.3–7.0) **(Table 1)**

### LV–LA Health phenotype distribution

LV-LA health phenotype distribution differed by indexation method. Using BSA-indexed LVM and LAV values, 2,899 participants were classified as preserved LV–LA Health (83%), 554 as mildly impaired (15.8%), and 60 as moderately/severely impaired (1.7%) **(Table 1a)**. With height²·⁷ indexation for both LVM and LAV, 2,097 participants were classified as preserved (59.7%), 1,115 as mildly impaired (31.7%), and 301 as moderately/severely impaired (8.6%)**(Table 1b)**.

Across the additional descriptive indexation strategies, preserved LV–LA Health ranged from 59.4% to 70.9%, mild impairment ranged from 25.6% to 33.3%, and moderate/severe impairment ranged from 3.4% to 8.5% **(Table 3).**

### Clinical and Echocardiographic Characteristics by Phenotype and Indexation Strategy

Clinical and echocardiographic characteristics varied across LV–LA Health phenotype categories under both indexation strategies (Tables 1a–2b). In the BSA-indexed analysis, older age, hypertension, smoking, and elevated NT-proBNP levels were more prevalent across worsening LV–LA Health phenotype categories (all P<.001). When phenotype was defined using height²·⁷ indexation, significant differences across phenotype categories were also observed for diabetes, dyslipidemia, and obesity (all P<.001).

In the BSA-indexed analysis, LAV, LAVi, LV mass, and LVMi all increased progressively across worsening phenotype categories (all P<.001). Similar patterns were observed with height²·⁷ indexation (P<.001). Mean LVEF remained within the normal range across phenotype categories and did not differ significantly by phenotype under either indexation strategy.

Abnormal GLS prevalence increased across phenotype categories under both strategies, although decreases in aabsolute GLS was only statistically significant in the BSA-indexed analysis.

### Impact of indexation strategy and reclassification

Compared with BSA indexation, height²·⁷ indexation resulted in a greater proportion of participants being classified as mildly impaired or moderately/severely impaired and was associated with different distributions of cardiometabolic risk factors across phenotype categories(Tables 1a–1b). Among participants classified as preserved, mean BMI was lower under height-based indexation (31 kg/m²) compared with BSA indexation (33 kg/m²), with corresponding obesity prevalence of 46% versus 57%. The magnitude of increase in BMI across phenotype categories was also greater under height-based indexation (31 to 40.9 kg/m²) compared with BSA indexation (32.7 to 35.2 kg/m²), with a corresponding increase in obesity prevalence from 46% to 90% under height-based scaling versus 57% to 68% under BSA indexation (both P<.001).

The prevalence of AHA/ACC-defined hypertension was 90.0% in the moderate/severe phenotype category under height²·⁷ indexation compared with 85.0% under BSA indexation (P<.001). Diabetes exhibited a non-monotonic pattern under BSA indexation but increased progressively across phenotype categories under height-based indexation (20.4% to 32.6%, P<.001). Dyslipidemia, family history, and total cardiovascular risk score differed across phenotype categories under both indexation strategies. Under height²·⁷ indexation, 17.2% of participants in the moderate/severe phenotype category had NT-proBNP ≥125 pg/mL compared with 13.6% under BSA indexation.

The proportion of Black participants increased across phenotype categories under both strategies, with greater concentration in the moderate/severe group under height-based indexation (59.5% vs. 56.7%). Additionally, sex differences were observed under height-based indexation but not under BSA indexation (P<.001 vs. P=0.058).

Reclassification analyses demonstrated changes in phenotype classification compared with the BSA-based reference, with all height-based indexation strategies resulting in reclassification toward higher severity (Table 5). Overall re-classification was 28.4% for height²·⁷ indexation compared to BSA indexation, with 28% of people being reclassified to a higher severity, and 1% to a lower severity.

### Supplemental GLS analysis

GLS was incorporated as a supplemental component to further stratify moderate/severe impairment in BSA indexation (Table 4). Among the 2,271 participants with available GLS data, incorporation of GLS into the BSA-indexed LV–LA Health phenotype resulted in 1,858 participants classified as preserved (81.81%), 348 as mildly impaired (15.32%), 62 as moderately impaired (2.73%), and 3 as severely impaired (0.13%). GLS was missing in 1,242 participants.

### Associations with LV–LA Health Phenotype using BSA Indexation

In multinomial regression analyses using BSA-indexed LV–LA Health phenotype with preserved as the reference group, several clinical variables were associated with worse phenotype categories (Tables 6A1–6A3). In univariable analyses (Table 6A1), older age, hypertension, current smoking, Black race, and higher BMI were associated with both mildly impaired and moderate/severely impaired LV–LA Health. The highest increased odds for moderate/severe impairment included Age per 10-year increase (OR 2.31; 95% CI, 1.69–3.14), hypertension (OR 2.84; 95% CI, 1.39–5.78), and current smoking (OR 2.20; 95% CI, 1.24–3.89).

After adjustment for age, several associations were attenuated compared with univariable analyses (Tables 6a1–6a2). Hypertension remained associated with mild impairment though effect attenuated (OR 1.96 vs 2.36) and was not statistically significant for increased odds of moderate/severe phenotype. Associations for BMI were largely unchanged after age adjustment. Current smoking remained associated with both mild and moderate/severe, though odds decreased. Black race also remained associated with mild (OR 1.57 vs. 1.68) and moderate/severe impairments (OR 1.86 vs. 2.12). Diabetes was no longer associated with moderate/severe impairment when adjusting for age.

In fully adjusted models, age, current smoking, and BMI remained associated with both mildly impaired and moderate/severely impaired LV–LA Health (Table 6a3). Age per 10-year increase was associated with moderate/severe impairment (OR 2.35; 95% CI, 1.69–3.26), and BMI remained associated with both mild (OR 1.07; 95% CI, 1.05–1.10) and moderate/severe impairment (OR 1.08; 95% CI, 1.01–1.15). Hypertension and Black race were associated with mild impairment but not moderate/severe impairment, while diabetes, dyslipidemia, family history, and high-risk county residence were not significantly associated.

### Associations with LV–LA Health Phenotype using Height²·⁷ Indexation

In multinomial regression analyses using height²·⁷-indexed LV–LA Health phenotype with preserved as the reference group, several clinical variables were associated with worse phenotype categories **(Tables 6b1–6b3)**. In univariable analyses **(Table 6b1)**, older age, hypertension, Black race, female sex, and higher BMI were associated with both mildly impaired and moderate/severely impaired LV–LA Health. The highest increased odds for moderate/severe impairment included obesity (OR 10.16; 95% CI, 6.94–14.88) and hypertension (OR 5.28; 95% CI, 3.59–7.79).

After adjustment for age, several associations were attenuated compared with univariable analyses (Tables 6b1–6b2). Hypertension remained strongly associated LV-LA health impairment though with decreased odds compared to the unadjusted model for both mild (OR 1.71 vs. 1.84) and moderate/severe (OR 4.58 vs. 5.28). BMI demonstrated minimal attenuation and remained strongly associated with both mild (OR 1.19 vs. 1.18) and moderate/severe impairment (OR 1.42 vs. 1.40 per kg/m²). Obesity had increased odds in the age adjusted model for both mild (OR 3.55 vs. 3.43) and moderate/severe impairments (OR 10.89 vs. 10.16).

In fully adjusted models, age, BMI, hypertension, current smoking, and female sex were associated with both mildly impaired and moderate/severely impaired LV–LA Health (Table 6B3). BMI demonstrated strong associations with both mild (OR 1.18; 95% CI, 1.16–1.21) and moderate/severe impairment (OR 1.39; 95% CI, 1.32–1.46). Hypertension remained associated with both mild (OR 1.24; 95% CI, 1.02–1.49) and moderate/severe impairment (OR 2.50; 95% CI, 1.65–3.79). Current smoking was associated with both mild (OR 1.29; 95% CI, 1.04–1.61) and moderate/severe impairment (OR 2.09; 95% CI, 1.49–2.93). Female sex was associated with both mild (OR 1.87; 95% CI, 1.57–2.23) and moderate/severe impairment (OR 1.64; 95% CI, 1.21–2.21). Black race was not associated with mild impairment but remained associated with moderate/severe impairment (OR 1.43; 95% CI, 1.09–1.88). Diabetes demonstrated an inverse association with mild impairment and was not associated with moderate/severe impairment, while dyslipidemia, family history, and high-risk county residence were not significantly associated with either mild or moderate/severe impairments.

### Biomarker Associations with LV–LA Health

In the biomarker substudy (N=970), distributions of NT-proBNP and troponin differed across LV–LA Health phenotype categories under both BSA- and height-indexed approaches (Tables 1a-1b). Under BSA indexation, the prevalence of NT-proBNP ≥125 pg/mL was 6.5% in the preserved group, 18.1% in the mildly impaired group, and 13.6% in the moderate/severe group (P<.001). Under height²·⁷ indexation, the prevalence increased from 6.7% to 8.9% to 17.2% across phenotype categories (P<.001). Troponin distributions also differed significantly across phenotype categories under both indexation strategies.

In adjusted biomarker analyses, elevated NT-proBNP was associated with mildly impaired LV–LA Health in the BSA-indexed phenotype (OR 2.10; 95% CI 1.19–3.70; P=0.011). Significant associations between NT-proBNP and moderate/severe impairment were observed in some models incorporating height-indexed LA volume, including LVM/BSA + LAV/height²·⁷ (OR 2.31; 95% CI 1.03–5.15; P=0.041). Most other NT-proBNP and troponin associations were not statistically significant (Table 7).

## Discussion

In this large community-based rural cohort, we developed and applied a composite LV–LA Health phenotype integrating left ventricular mass and left atrial volume to characterize subclinical structural cardiac remodeling. Several key findings emerged. First, construction of the phenotype was feasible in 92% of participants, supporting its potential applicability in large-scale community-based echocardiographic studies. Second, LV–LA Health phenotype severity demonstrated graded associations with cardiometabolic risk factors and echocardiographic measures of remodeling, supporting the construct validity of the phenotype (Tables 1a–2b).

Third, phenotype distribution differed substantially across indexation strategies, with height-based approaches classifying a greater proportion of participants as having impaired LV–LA Health. Finally, age, body mass index, hypertension, and smoking demonstrated consistent associations with worsening phenotype severity across regression models, although the magnitude and significance of some associations differed according to indexation strategy.

### LV–LA Health as a Composite Marker of Subclinical Remodeling

Our findings support the concept that integrating LV and LA structural measures captures a broader spectrum of subclinical cardiac remodeling than either metric alone. LVMi reflects myocardial adaptation to chronic pressure and volume loading, whereas LAVi reflects cumulative diastolic burden and chronically elevated filling pressures (8,9). The graded associations observed across phenotype categories with both clinical risk factors and echocardiographic measures suggest that this composite phenotype represents a biologically coherent continuum of cardiac remodeling. Prior studies have demonstrated that abnormalities in LV mass and LA volume independently predict adverse cardiovascular outcomes, including heart failure, atrial fibrillation, and mortality (16–19). The present findings extend this work by demonstrating that a composite phenotype incorporating both domains captures clinically meaningful variation in cardiometabolic risk and structural remodeling.

### Impact of Indexation Strategy

A central finding of this study is that LV–LA Health phenotype classification varied substantially according to indexation strategy. Compared with BSA indexation, height-based approaches identified a greater proportion of participants with impaired LV–LA Health and produced more pronounced gradients across cardiometabolic risk factors and echocardiographic measures **(Tables 1A–2B, Table 3)**. In particular, obesity and diabetes demonstrated more consistent increases across phenotype severity categories under height-based scaling **(Table 1B)**.

These findings likely reflect fundamental differences in how body size is accounted for by each indexing method. Because BSA incorporates weight, it may partially normalize cardiac enlargement associated with excess adiposity. In contrast, height-based indexation was associated with stronger relationships between phenotype severity and measures of obesity, hypertension, and structural remodeling within this cohort **(Tables 1A–2B)**.

Height-based indexation also resulted in substantial upward reclassification of phenotype severity compared with the BSA reference **(Table 5)**. Participants reclassified into more severe categories generally exhibited greater cardiometabolic burden and less favorable echocardiographic profiles, suggesting that alternative indexation strategies identify meaningful differences in structural remodeling. Whether these differences translate into improved prognostic performance requires longitudinal outcome validation.

### Determinants of LV–LA Health (Regression Findings)

To further understand the determinants of LV–LA Health phenotype, we evaluated associations using multinomial regression models. Across both indexation strategies, BMI, hypertension, and age emerged as consistent determinants of LV–LA Health phenotype. Notably, BMI remained strongly and independently associated with both mild and moderate/severe impairment even after multivariable adjustment, with substantially greater effect sizes observed under height-based indexation. This finding reinforces the central role of adiposity in cardiac remodeling and suggests that BSA indexation may attenuate the relationship between obesity and cardiac structure.

Hypertension also demonstrated consistent associations with LV–LA Health, reflecting its established role in driving both LV hypertrophy and LA enlargement. However, important differences were observed across indexation strategies. Hypertension remained associated with both mild and moderate/severe impairment under height-based models but was attenuated for moderate/severe impairment under BSA indexation.

Smoking remained associated with worse phenotype categories in adjusted models despite less consistent patterns in descriptive analyses, suggesting the presence of confounding factors such as age (Tables 1A–1B, Tables 6A3–6B3).

Associations with sex and race varied according to indexation strategy, underscoring the importance of considering scaling methodology when interpreting demographic differences in cardiac structure (Tables 6A3 and 6B3)..

### Biomarker Associations and Construct Validity

In descriptive analyses, worsening LV–LA Health phenotype severity was associated with less favorable NT-proBNP and troponin distributions under both BSA- and height-based indexation strategies, consistent with increasing myocardial stress and structural remodeling (Tables 1A–1B). However, adjusted biomarker associations were less consistent. Elevated NT-proBNP remained associated with impaired phenotype in selected models, whereas many other NT-proBNP and troponin associations were not statistically significant (Table 7).

These findings provide limited supportive evidence that the LV–LA Health phenotype reflects aspects of subclinical structural remodeling associated with myocardial stress. However, the variability of biomarker associations across models suggests that the relationship between LV–LA Health phenotype, circulating biomarkers, and cardiovascular risk is complex and requires further investigation in larger cohorts with longitudinal follow-up (Table 7).

## Limitations

Several limitations should be considered in this study. The cross-sectional design limits causal inference and assessment of temporal relationships between cardiometabolic risk factors and structural remodeling. The interpretation of height-based indexation is limited by the absence of widely validated reference ranges. The abnormal thresholds used for height-based indexation were derived from prior investigations performed in predominantly non-obese populations outside the United States. As a result, the applicability of these cut points to a rural U.S. population with a high prevalence of obesity remains uncertain. As a result, although height-based indexation demonstrated stronger associations with cardiometabolic risk factors and more consistent gradient patterns, it remains unclear whether these thresholds optimally define normal versus abnormal cardiac structure. This limitation is particularly relevant when interpreting the observed reclassification, as shifts in phenotype severity may reflect differences in scaling rather than true differences in pathologic remodeling.

Additionally, longitudinal data is needed to determine whether differences in LV–LA Health phenotypes translate into differences in clinical outcomes or risks. Although both LV mass and LA volume are established predictors of cardiovascular events (16–19), the prognostic implications of the composite LV–LA Health phenotype requires prospective validation

Selection and ascertainment biases could also be present. Participants included in the analytic cohort required successful acquisition of echocardiographic measurements for both LV mass and LA volume, and those with missing data were excluded. Although feasibility was high, this may preferentially exclude individuals with poorer image quality or more advanced comorbidity.

Furthermore, the RURAL cohort represents a geographically and demographically specific population, which may limit generalizability to other populations with different risk profiles or healthcare access.

Residual confounding remains possible despite multivariable adjustment. Behavioral and socioeconomic factors such as access to care, health literacy, and longitudinal treatment patterns, may influence both cardiometabolic risk factor burden and cardiac remodeling but were not fully captured. These factors could possibly impact observed associations across indexation strategies.

Measurement-related limitations should also be considered. Although echocardiographic acquisition followed standardized protocols with AI assistance, variability in image quality and reliance on geometric assumptions for LV mass and LA volume estimation may introduce measurement error.

## Future Directions

Future studies should extend these findings by incorporating longitudinal follow-up of clinical outcomes to evaluate the prognostic significance of the LV–LA Health phenotype and to evaluate whether height-based indexation provides incremental prognostic information beyond conventional BSA-based approaches. In addition, there is a need to establish and validate normative reference values for height-indexed LV mass and LA volume across diverse populations, particularly in cohorts with a high prevalence of obesity.

## Conclusion

The LV–LA Health phenotype represents a feasible and biologically coherent measure of subclinical cardiac remodeling. Indexation strategy significantly influences phenotype classification and its clinical associations, with height-based indexation identifying a greater burden of impairment and demonstrating stronger relationships with cardiometabolic risk factors. These findings highlight the importance of scaling methodology in echocardiographic assessment and support further investigation of height-based indexation in cardiovascular risk stratification and outcome prediction.

## 4. Statistical Results

**Table 0.** Summary data for Inclusion / Exclusion Criteria

| **Inclusion / Exclusion Criteria** | **N remaining** | **N excluded (%)** |
| --- | --- | --- |
| Starting Population:<br>RURAL ECHO cohort<br>with an attempted<br>echocardiographic exam | 3833 |  |
| **Patient Exclusion**<br>**Criteria:** |  |  |
| Exclude Patients with<br>missing data for LA<br>volume | 3732 | 101(2.6%) |
| Exclude Patients with<br>missing data for LV mass | 3513 | 219 (5.8%) |
| Exclude Patients with<br>missing data for BSA and<br>Height<br>**(= Main Population)** | 3513 | 0 |
| Exclude Patients with<br>missing data for GLS<br>**(=Supplementary**<br>**Population)** | 2271 | 1242(35.3%) |

\*\*\*The proportion of RURAL ECHO participants with a feasible LV-LA Health Score is 92 % (3513/3833) .

**Table 1A.**
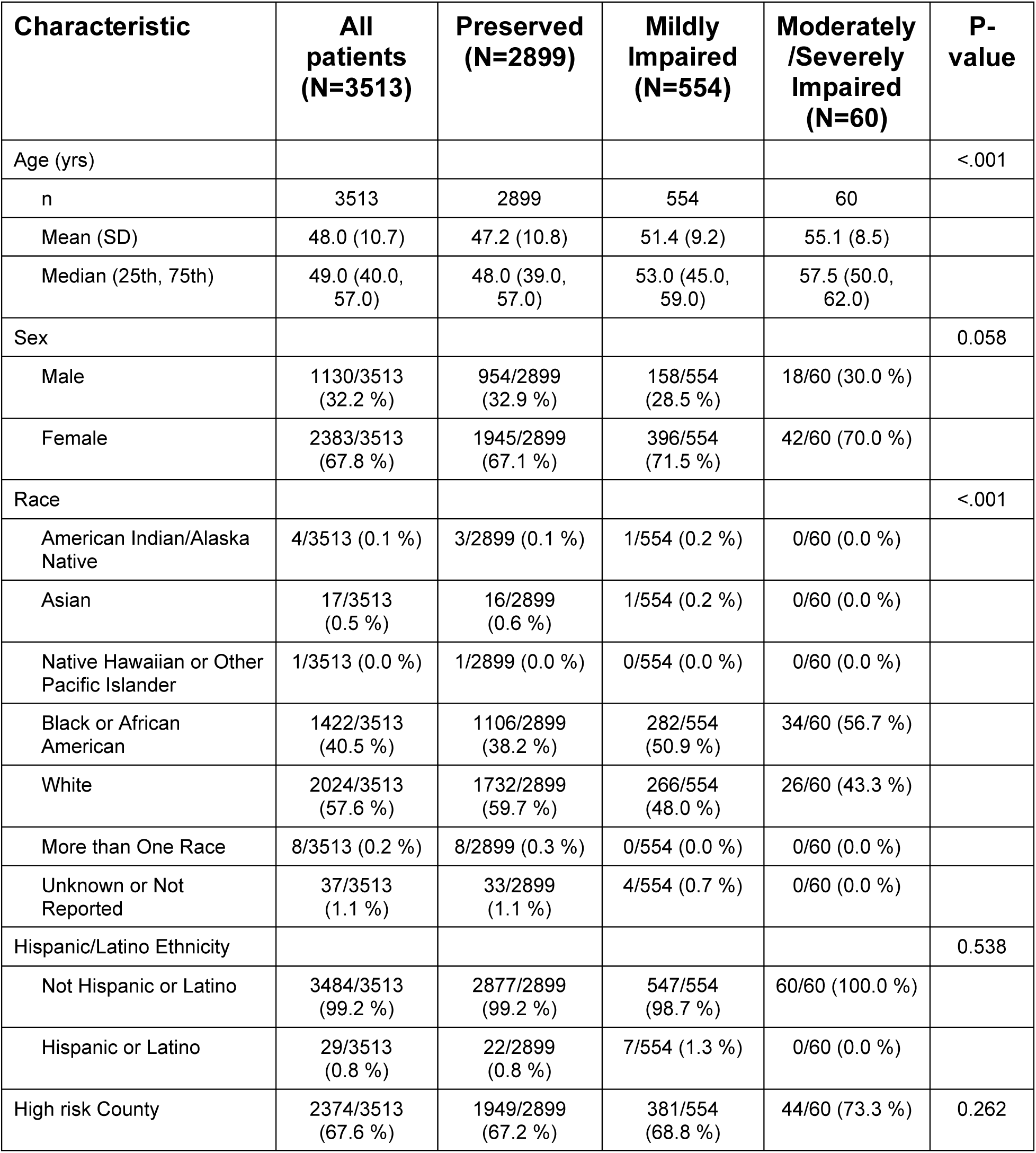
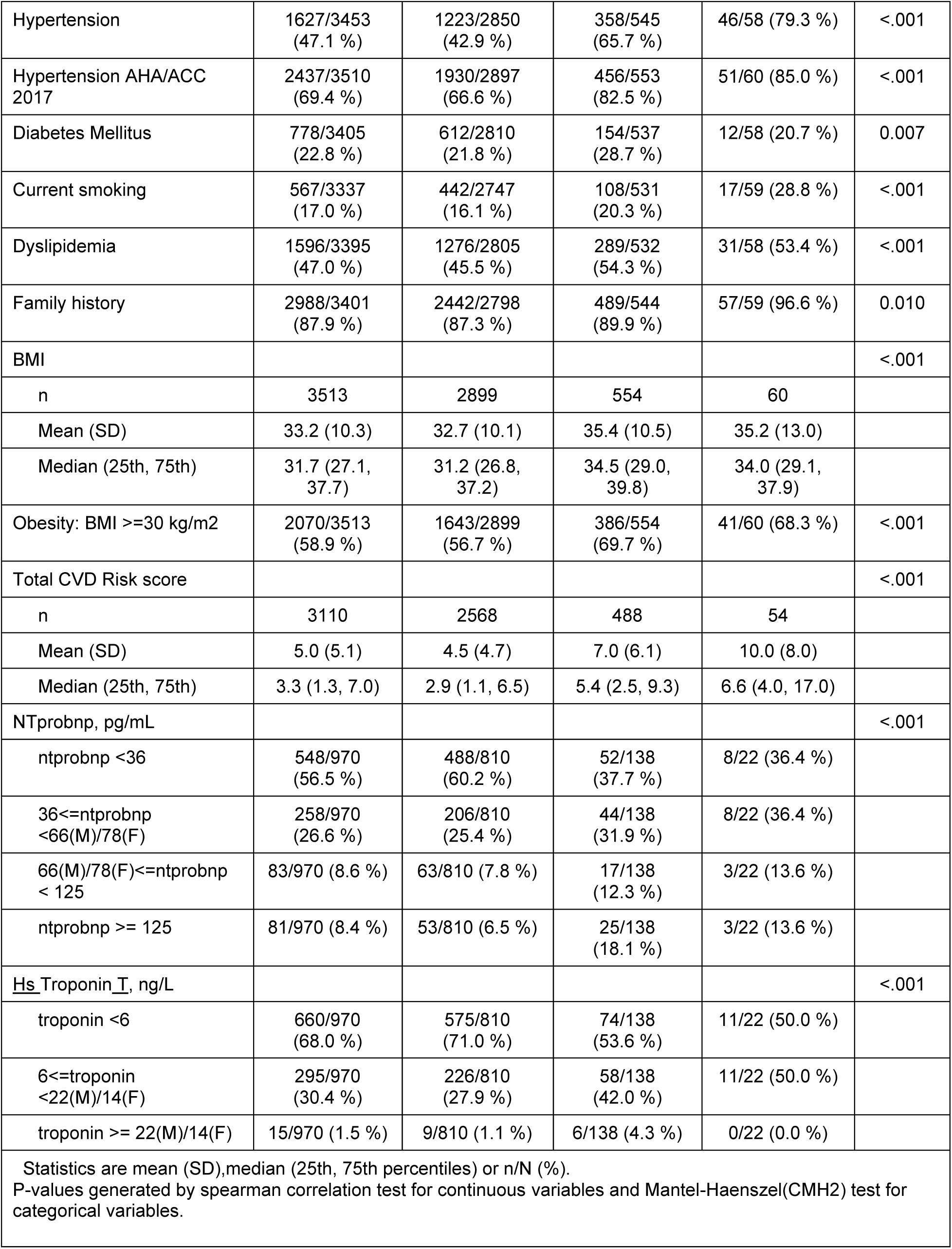
Demographic Characteristics by LA-LV Health Score Category based on BSA-indexed variables <u>for both LA and LV</u>

**Table 1B.** Demographic Characteristics by LA-LV Health Score Category based on height-indexed variable: Primary Endpoint 3: LVLA Health Phenotype (both height-indexed ^2.7 & ^2.7)

| **Characteristic** | **All patients**<br>**(N=3513)** | **Preserved**<br>**(N=2097)** | **Mildly**<br>**Impaired**<br>**(N=1115)** | **Moderately**<br>**/Severely**<br>**Impaired**<br>**(N=301)** | **P-**<br>**value** |
| --- | --- | --- | --- | --- | --- |
| Age (yrs) |  |  |  |  | <.001 |
| n | 3513 | 2097 | 1115 | 301 |  |
| Mean (SD) | 48.0 (10.7) | 47.0 (10.9) | 49.0 (10.2) | 51.1 (10.1) |  |
| Median (25th, 75th) | 49.0 (40.0, 57.0) | 47.0 (38.0, 56.0) | 50.0 (42.0, 58.0) | 53.0 (45.0, 59.0) |  |
| Sex |  |  |  |  | <.001 |
| Male | 1130/3513<br>(32.2 %) | 777/2097<br>(37.1 %) | 277/1115<br>(24.8 %) | 76/301 (25.2 %) |  |
| Female | 2383/3513<br>(67.8 %) | 1320/2097<br>(62.9 %) | 838/1115<br>(75.2 %) | 225/301 (74.8 %) |  |
| Race |  |  |  |  | <.001 |
| American Indian/Alaska Native | 4/3513 (0.1 %) | 3/2097 (0.1 %) | 0/1115 (0.0 %) | 1/301 (0.3 %) |  |
| Asian | 17/3513 (0.5 %) | 14/2097 (0.7 %) | 3/1115 (0.3 %) | 0/301 (0.0 %) |  |
| Native Hawaiian or Other Pacific Islander | 1/3513 (0.0 %) | 0/2097 (0.0 %) | 0/1115 (0.0 %) | 1/301 (0.3 %) |  |
| Black or African American | 1422/3513<br>(40.5 %) | 736/2097<br>(35.1 %) | 507/1115<br>(45.5 %) | 179/301 (59.5 %) |  |
| White | 2024/3513<br>(57.6 %) | 1311/2097<br>(62.5 %) | 595/1115<br>(53.4 %) | 118/301 (39.2 %) |  |
| More than One Race | 8/3513 (0.2 %) | 6/2097 (0.3 %) | 1/1115 (0.1 %) | 1/301 (0.3 %) |  |
| Unknown or Not Reported | 37/3513 (1.1 %) | 27/2097 (1.3 %) | 9/1115 (0.8 %) | 1/301 (0.3 %) |  |
| Hispanic/Latino Ethnicity |  |  |  |  | 0.166 |
| Not Hispanic or Latino | 3484/3513<br>(99.2 %) | 2084/2097<br>(99.4 %) | 1102/1115<br>(98.8 %) | 298/301 (99.0 %) |  |
| Hispanic or Latino | 29/3513 (0.8 %) | 13/2097 (0.6 %) | 13/1115 (1.2 %) | 3/301 (1.0 %) |  |
| High risk County | 2374/3513<br>(67.6 %) | 1392/2097<br>(66.4 %) | 759/1115<br>(68.1 %) | 223/301 (74.1 %) | 0.013 |
| Hypertension | 1627/3453<br>(47.1 %) | 807/2064<br>(39.1 %) | 594/1094<br>(54.3 %) | 226/295 (76.6 %) | <.001 |
| Hypertension AHA/ACC 2017 | 2437/3510<br>(69.4 %) | 1322/2096<br>(63.1 %) | 846/1115<br>(75.9 %) | 269/299 (90.0 %) | <.001 |
| Diabetes Mellitus | 778/3405<br>(22.8 %) | 414/2033<br>(20.4 %) | 269/1081<br>(24.9 %) | 95/291 (32.6 %) | <.001 |
| Current smoking | 567/3337<br>(17.0 %) | 331/1977<br>(16.7 %) | 175/1069<br>(16.4 %) | 61/291 (21.0 %) | 0.244 |
| Dyslipidemia | 1596/3395<br>(47.0 %) | 885/2034<br>(43.5 %) | 557/1071<br>(52.0 %) | 154/290 (53.1 %) | <.001 |
| Family history | 2988/3401<br>(87.9 %) | 1750/2019<br>(86.7 %) | 961/1085<br>(88.6 %) | 277/297 (93.3 %) | 0.001 |
| BMI |  |  |  |  | <.001 |
| n | 3513 | 2097 | 1115 | 301 |  |
| Mean (SD) | 33.2 (10.3) | 30.7 (10.1) | 35.6 (8.0) | 40.9 (13.0) |  |
| Median (25th, 75th) | 31.7 (27.1, 37.7) | 29.4 (25.5, 34.1) | 34.8 (30.0, 40.0) | 39.2 (34.4, 44.6) |  |
| Obesity: BMI >=30 kg/m2 | 2070/3513<br>(58.9 %) | 968/2097<br>(46.2 %) | 832/1115<br>(74.6 %) | 270/301 (89.7 %) | <.001 |
| Total CVD Risk score |  |  |  |  | <.001 |
| n | 3110 | 1856 | 990 | 264 |  |
| Mean (SD) | 5.0 (5.1) | 4.3 (4.7) | 5.5 (5.3) | 7.6 (6.7) |  |
| Median (25th, 75th) | 3.3 (1.3, 7.0) | 2.7 (1.0, 6.3) | 3.9 (1.6, 7.6) | 5.9 (2.8, 10.2) |  |
| NTprobnp, pg/mL |  |  |  |  | <.001 |
| ntprobnp <36 | 548/970 (56.5 %) | 366/597 (61.3 %) | 145/280 (51.8 %) | 37/93 (39.8 %) |  |
| 36<=ntprobnp<br><66(M)/78(F) | 258/970 (26.6 %) | 149/597 (25.0 %) | 80/280 (28.6 %) | 29/93 (31.2 %) |  |
| 66(M)/78(F)<=ntprobnp <<br>125 | 83/970 (8.6 %) | 42/597 (7.0 %) | 30/280 (10.7 %) | 11/93 (11.8 %) |  |
| ntprobnp >= 125 | 81/970 (8.4 %) | 40/597 (6.7 %) | 25/280 (8.9 %) | 16/93 (17.2 %) |  |
| Troponin, ng/L |  |  |  |  | 0.004 |
| troponin <6 | 660/970 (68.0 %) | 426/597 (71.4 %) | 183/280 (65.4 %) | 51/93 (54.8 %) |  |
| 6<=troponin <22(M)/14(F) | 295/970 (30.4 %) | 165/597 (27.6 %) | 89/280 (31.8 %) | 41/93 (44.1 %) |  |
| troponin >= 22(M)/14(F) | 15/970 (1.5 %) | 6/597 (1.0 %) | 8/280 (2.9 %) | 1/93 (1.1 %) |  |
| Statistics are mean (SD),median (25th, 75th percentiles) or n/N (%).<br>P-values generated by spearman correlation test for continuous variables and Mantel-Haenszel(CMH2) test for categorical variables. |  |  |  |  |  |

**Table 2A.** Echocardiographic Parameter by LA-LV Health Score Category based on BSA-indexed <u>both</u> variables

| **Characteristic** | **All patients**<br>**(N=3513)** | **Preserved**<br>**(N=2899)** | **Mildly**<br>**Impaired**<br>**(N=554)** | **Moderately**<br>**/Severely**<br>**Impaired**<br>**(N=60)** | **P-**<br>**value** |
| --- | --- | --- | --- | --- | --- |
| BSA, Men |  |  |  |  | 0.040 |
| N | 1130 | 954 | 158 | 18 |  |
| Mean (SD) | 2.14 (0.24) | 2.14 (0.24) | 2.18 (0.27) | 2.06 (0.33) |  |
| Median (25th, 75th) | 2.13 (1.97-2.29) | 2.13 (1.97-2.28) | 2.18 (2.01-2.34) | 2.10 (1.87-2.26) |  |
| BSA, Women |  |  |  |  | <.001 |
| N | 2383 | 1945 | 396 | 42 |  |
| Mean (SD) | 1.92 (0.24) | 1.91 (0.25) | 1.95 (0.23) | 1.95 (0.18) |  |
| Median (25th, 75th) | 1.90 (1.75-2.07) | 1.89 (1.74-2.06) | 1.95 (1.79-2.09) | 1.96 (1.84-2.07) |  |
| BP and SP LV Ejection Fraction |  |  |  |  | 0.051 |
| N | 3498 | 2886 | 552 | 60 |  |
| Mean (SD) | 58.8 (5.0) | 59.1 (4.2) | 57.9 (7.2) | 55.9 (9.3) |  |
| Median (25th, 75th) | 60.0 (56.2-60.9) | 60.0 (56.5-60.9) | 60.0 (55.0-61.1) | 59.3 (55.0-60.0) |  |
| LVIDd Diameter: average |  |  |  |  | <.001 |
| N | 3513 | 2899 | 554 | 60 |  |
| Mean (SD) | 4.4 (0.5) | 4.4 (0.5) | 4.6 (0.6) | 5.0 (0.6) |  |
| Median (25th, 75th) | 4.4 (4.1-4.8) | 4.4 (4.1-4.7) | 4.6 (4.2-4.9) | 5.0 (4.7-5.4) |  |
| LA Systolic Volume |  |  |  |  | <.001 |
| N | 3513 | 2899 | 554 | 60 |  |
| Mean (SD) | 46.7 (18.6) | 42.8 (15.1) | 63.6 (21.5) | 82.9 (20.2) |  |
| Median (25th, 75th) | 46.0 (34.5-57.7) | 43.7 (32.6-53.6) | 65.7 (47.7-77.2) | 78.6 (72.2-89.9) |  |
| LAVi BSA |  |  |  |  | <.001 |
| N | 3513 | 2899 | 554 | 60 |  |
| Mean (SD) | 23.4 (8.8) | 21.5 (7.0) | 31.6 (10.1) | 41.8 (9.1) |  |
| Median (25th, 75th) | 23.4 (17.9-28.6) | 22.3 (17.1-26.8) | 34.4 (24.2-38.0) | 39.6 (36.9-43.3) |  |
| LAVi BSA, elevated |  |  |  |  | <.001 |
| Normal | 3162/3513<br>(90.0 %) | 2899/2899<br>(100.0 %) | 263/554<br>(47.5 %) | 0/60 (0.0 %) |  |
| Abnormal | 351/3513<br>(10.0 %) | 0/2899 (0.0 %) | 291/554<br>(52.5 %) | 60/60 (100.0 %) |  |
| LV mass, gms |  |  |  |  | <.001 |
| N | 3513 | 2899 | 554 | 60 |  |
| Mean (SD) | 153.7 (49.0) | 144.1 (39.4) | 195.2 (62.4) | 236.3 (53.0) |  |
| Median (25th, 75th) | 146.8 (119.6-178.8) | 140.1 (115.2-167.7) | 184.5 (151.4-230.8) | 228.0 (193.5-260.7) |  |
| LVMi BSA |  |  |  |  | <.001 |
| N | 3513 | 2899 | 554 | 60 |  |
| Mean (SD) | 76.7 (20.1) | 72.0 (15.0) | 96.2 (25.3) | 118.9 (21.9) |  |
| Median (25th, 75th) | 73.9 (63.1-87.1) | 71.1 (61.7-81.8) | 96.2 (76.8-109.7) | 111.5 (100.1-130.6) |  |
| LVMi BSA, elevated |  |  |  |  | <.001 |
| Normal | 3190/3513 (90.8 %) | 2899/2899 (100.0 %) | 291/554 (52.5 %) | 0/60 (0.0 %) |  |
| Abnormal | 323/3513 (9.2 %) | 0/2899 (0.0 %) | 263/554 (47.5 %) | 60/60 (100.0 %) |  |
| Global Longitudinal Strain-absolute |  |  |  |  | <.001 |
| N | 2271 | 1888 | 347 | 36 |  |
| Mean (SD) | 21.4 (3.3) | 21.7 (3.1) | 20.5 (4.1) | 19.2 (4.5) |  |
| Median (25th, 75th) | 21.6 (19.7-23.4) | 21.7 (19.9-23.4) | 20.8 (18.2-23.5) | 19.9 (17.1-22.1) |  |
| Global Longitudinal Strain-low |  |  |  |  | <.001 |
| Normal | 2209/2271 (97.3 %) | 1858/1888 (98.4 %) | 318/347 (91.6 %) | 33/36 (91.7 %) |  |
| Abnormal | 62/2271 (2.7 %) | 30/1888 (1.6 %) | 29/347 (8.4 %) | 3/36 (8.3 %) |  |
| Statistics are mean (SD),median (25th, 75th percentiles) or n/N (%).<br>P-values generated by spearman correlation test for continuous variables and Mantel-Haenszel(CMH2) test for categorical variables. |  |  |  |  |  |

**Table 2B.**
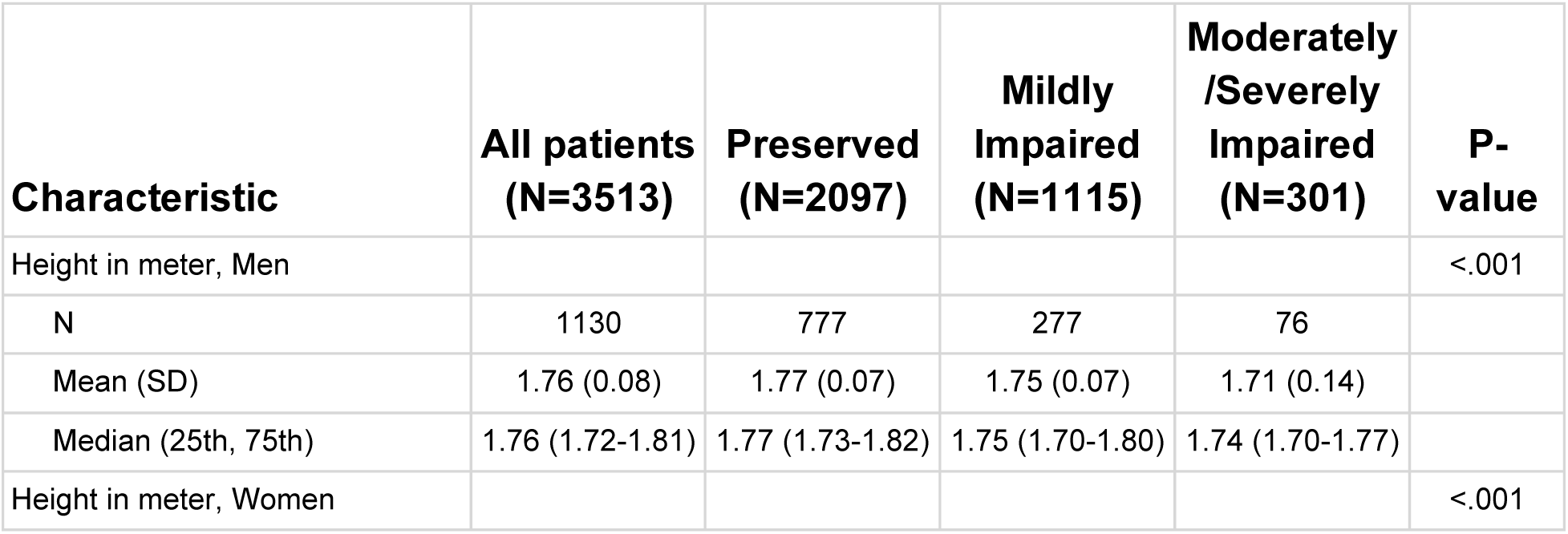
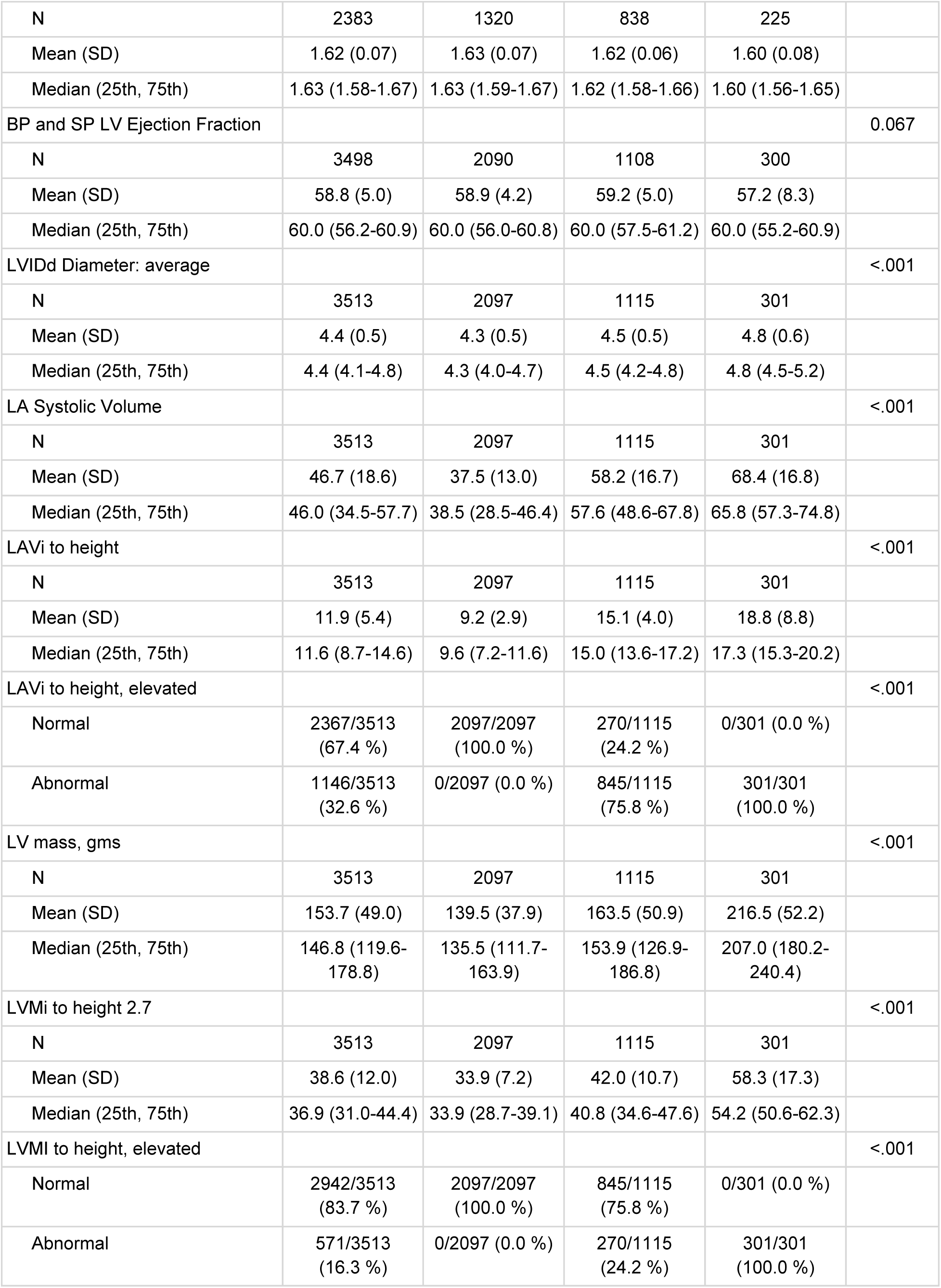
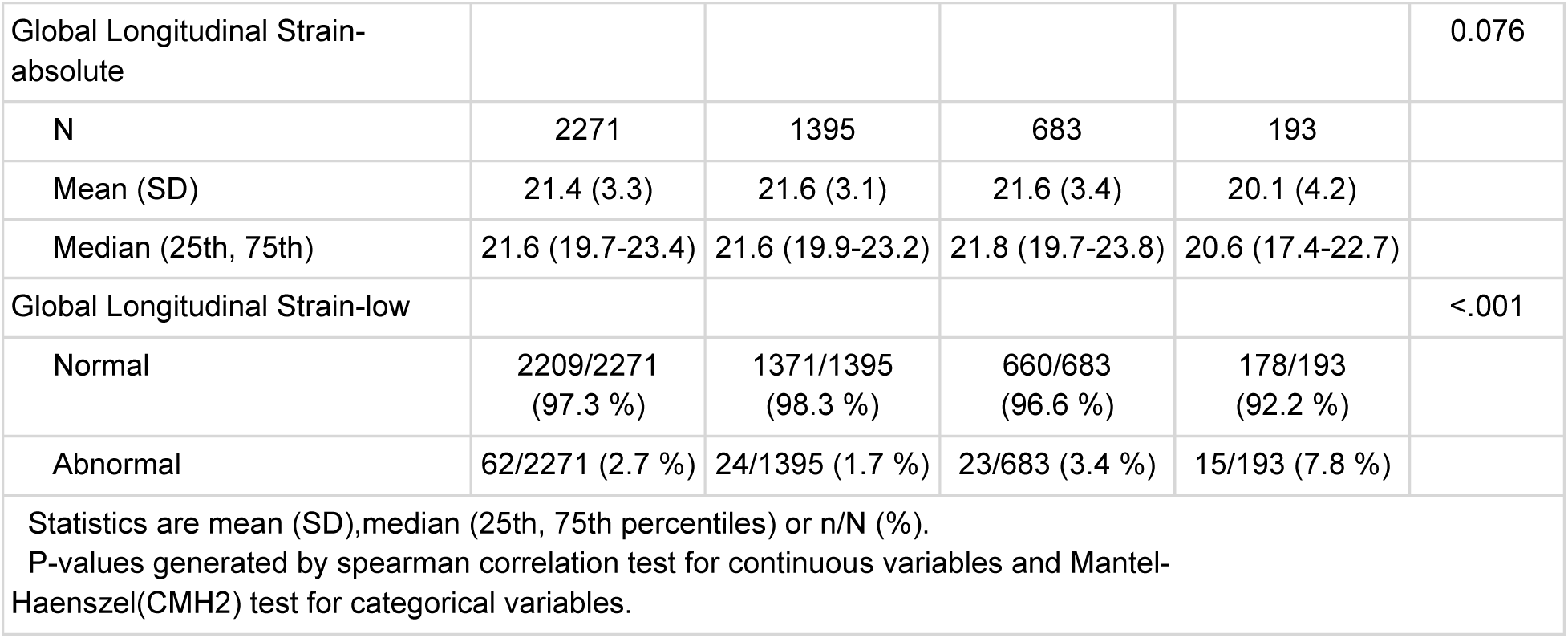
Echocardiographic Parameter by LA-LV Health Score Category based on height-indexed <u>both</u> variable: Primary Endpoint 3: LV–LA Health Phenotype (height-indexed ^2.7 & ^2.7)

**Table 3.** Distribution of LV LA Health Phenotypes by LAV and LVM Indexation Strategy in the RURAL Cohort

| *LV LA Health Phenotype* | *LVM/BSA and LAV/BSA* | *LVM/H<sup>2.7</sup> and LAV/H<sup>2.7</sup>* | *LVM/H<sup>2.7</sup> and LAV/H<sup>2</sup>* | *LVM/H<sup>2.7</sup> and LAV/H<sup>1.7</sup>* | *LVM/BSA and LAV/H<sup>2.7</sup>* | *LVM/BSA and LAV/H<sup>2</sup>* | *LVM/BSA and LAV/H<sup>1.7</sup>* |
| --- | --- | --- | --- | --- | --- | --- | --- |
| 0=Preserved (n, %) | 2899(82.5%) | 2097(59.7%) | 2361(67.2%) | 2086(59.4%) | 2197(62.5%) | 2492(70.9%) | 2189(62.3%) |
| 1=Mild impairment (n, %) | 554(15.8%) | 1115(31.7%) | 914(26.0%) | 1128(32.1%) | 1163(33.1%) | 900(25.6%) | 1170(33.3%) |
| 2=Moderate/severe impairment (n, %) | 60(1.7%) | 301(8.6%) | 238(6.8%) | 299(8.5%) | 153(4.4%) | 121(3.4%) | 154(4.4%) |
| Total N | 3513 | 3513 | 3513 | 3513 | 3513 | 3513 | 3513 |

**Legend: See Appendix for cutpoints based on different indexation strategies.**

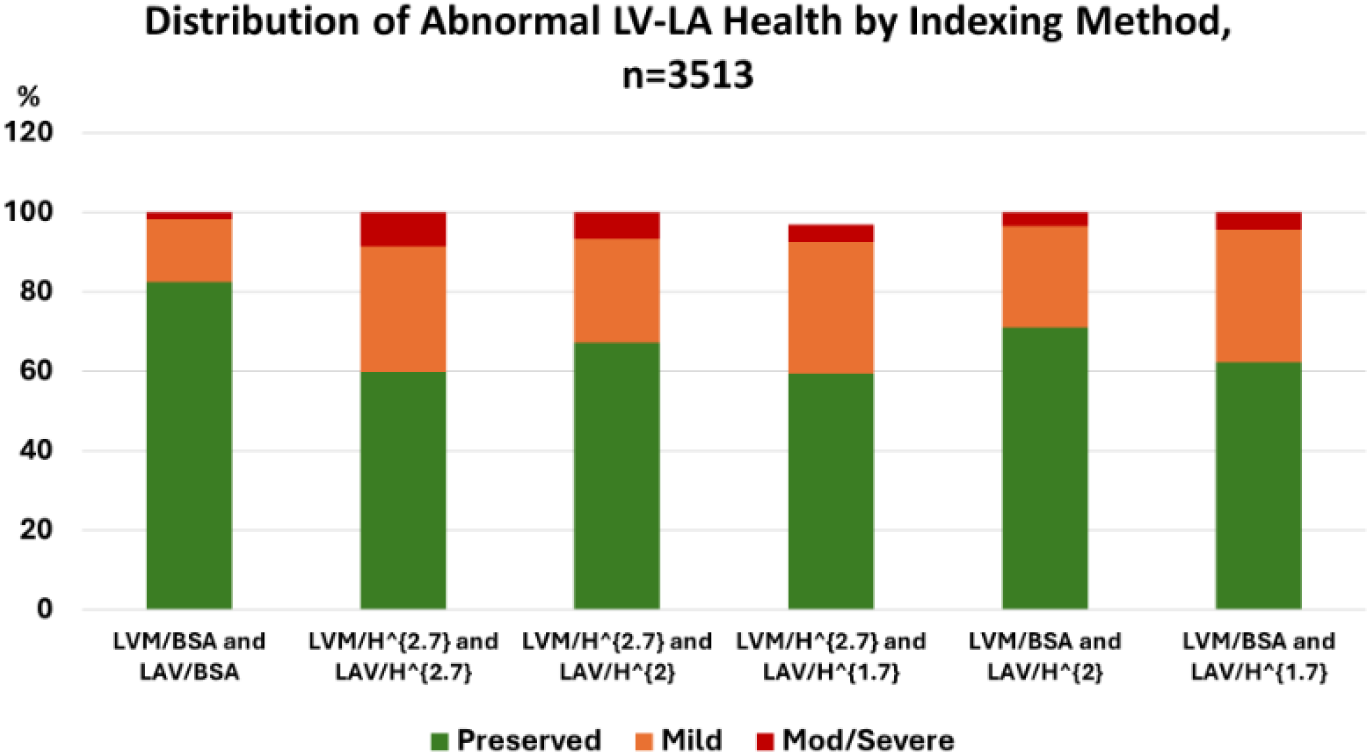

**Table 4.** LV-LA health distribution with addition of GLS

| *BSA-Indexed LV LA Health (with GLS)* |  |  |  |  |
| --- | --- | --- | --- | --- |
| *lvla_abnormal_s* | *Frequency* | *Percent* | *Cumulative Frequency* | *Cumulative Percent* |
| *Preserved* | 1858 | 81.81 | 1858 | 81.81 |
| *Mildly impaired* | 348 | 15.32 | 2206 | 97.14 |
| *Moderately impaired* | 62 | 2.73 | 2268 | 99.87 |
| *Severely impaired* | 3 | 0.13 | 2271 | 100.00 |
| *Frequency Missing = 1242* |  |  |  |  |

**Legend:** LVLA Health with incorporation of GLS. LVM and LAV were indexed to body surface area (BSA). See Appendix for abnormal cutpoints. GLS was not indexed with the absolute value taken. For GLS, abnormal defined as absolute value <15%.

**Table 5:**
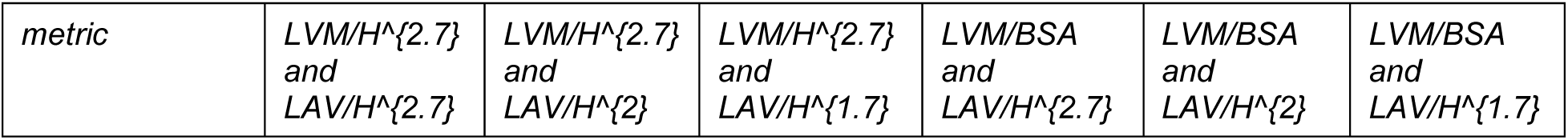
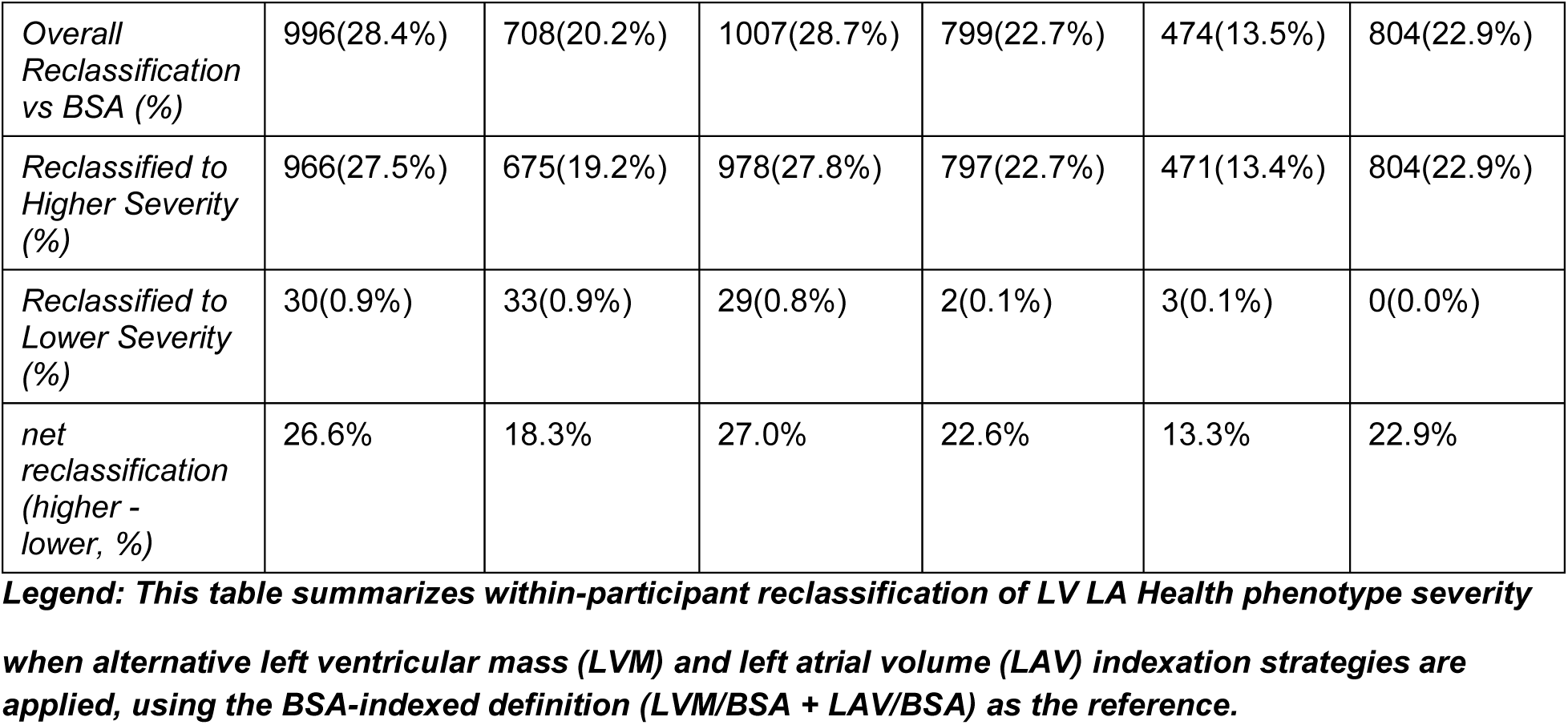
Reclassification of LV LA Health Phenotypes Across Indexation Strategies Relative to BSA Reference

| *metric* | *LVM/H<sup>2.7</sup> and LAV/H<sup>2.7</sup>* | *LVM/H<sup>2.7</sup> and LAV/H<sup>2</sup>* | *LVM/H<sup>2.7</sup> and LAV/H<sup>1.7</sup>* | *LVM/BSA and LAV/H<sup>2.7</sup>* | *LVM/BSA and LAV/H<sup>2</sup>* | *LVM/BSA and LAV/H<sup>1.7</sup>* |
| --- | --- | --- | --- | --- | --- | --- |
| *Overall*<br>*Reclassification*<br>*vs BSA (%)* | 996(28.4%) | 708(20.2%) | 1007(28.7%) | 799(22.7%) | 474(13.5%) | 804(22.9%) |
| *Reclassified to*<br>*Higher Severity*<br>*(%)* | 966(27.5%) | 675(19.2%) | 978(27.8%) | 797(22.7%) | 471(13.4%) | 804(22.9%) |
| *Reclassified to*<br>*Lower Severity*<br>*(%)* | 30(0.9%) | 33(0.9%) | 29(0.8%) | 2(0.1%) | 3(0.1%) | 0(0.0%) |
| *net*<br>*reclassification*<br>*(higher -*<br>*lower, %)* | 26.6% | 18.3% | 27.0% | 22.6% | 13.3% | 22.9% |

**Legend:** This table summarizes within-participant reclassification of LV LA Health phenotype severity when alternative left ventricular mass (LVM) and left atrial volume (LAV) indexation strategies are applied, using the BSA-indexed definition (LVM/BSA + LAV/BSA) as the reference.

**Table 6a1:**
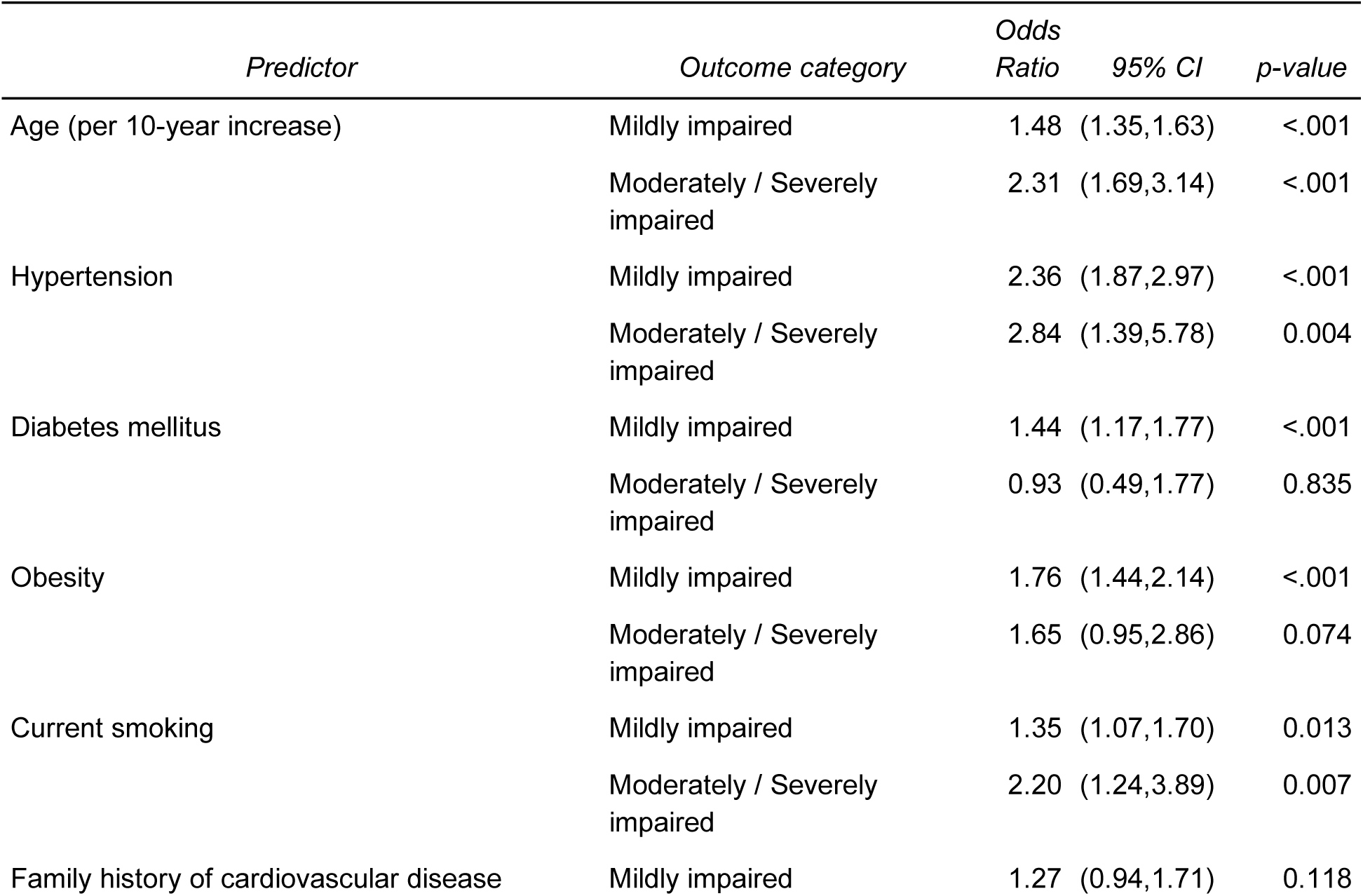
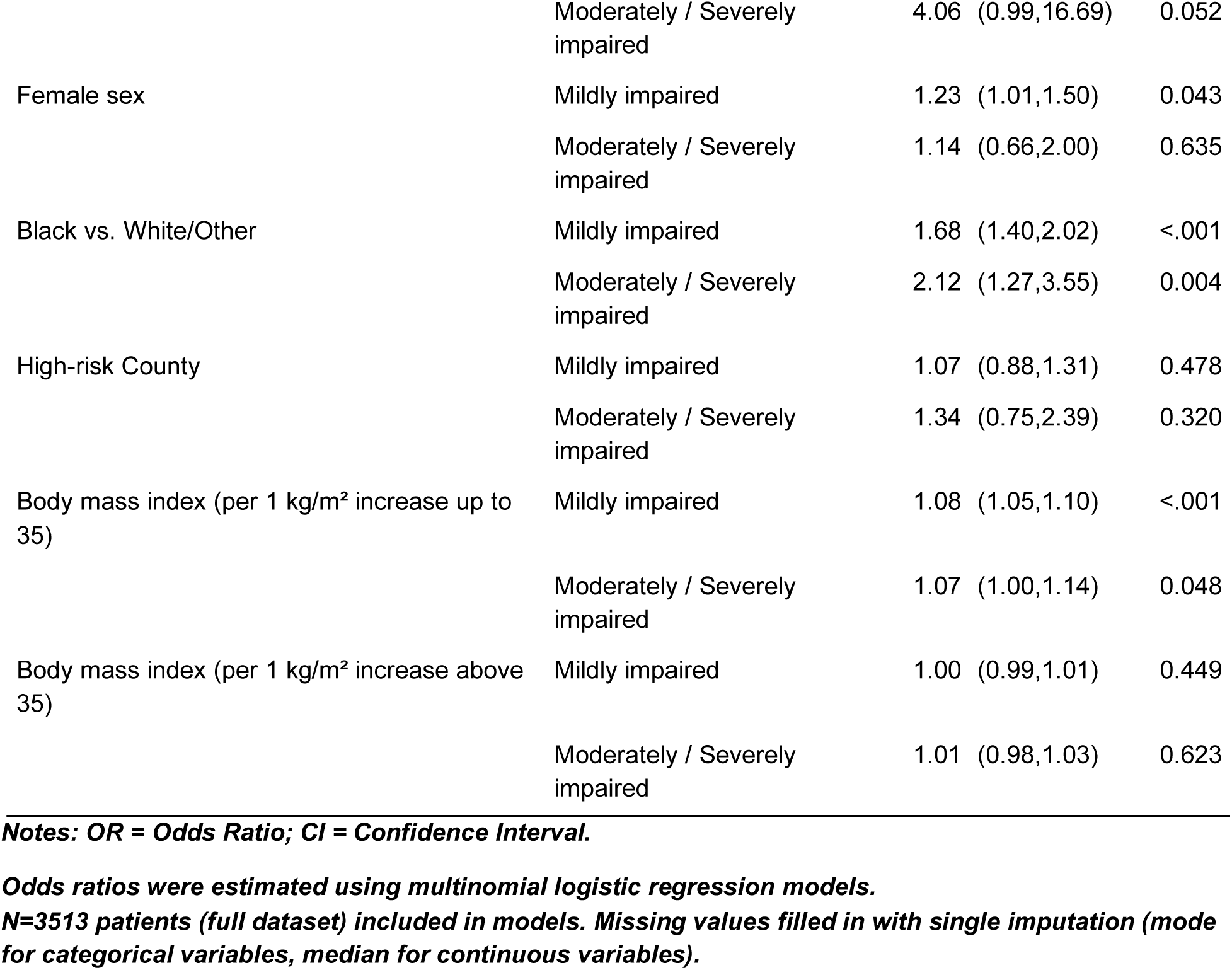
Univariable Multinomial Logistic Regression Results (Reference = Preserved)

**Table 6a2:**
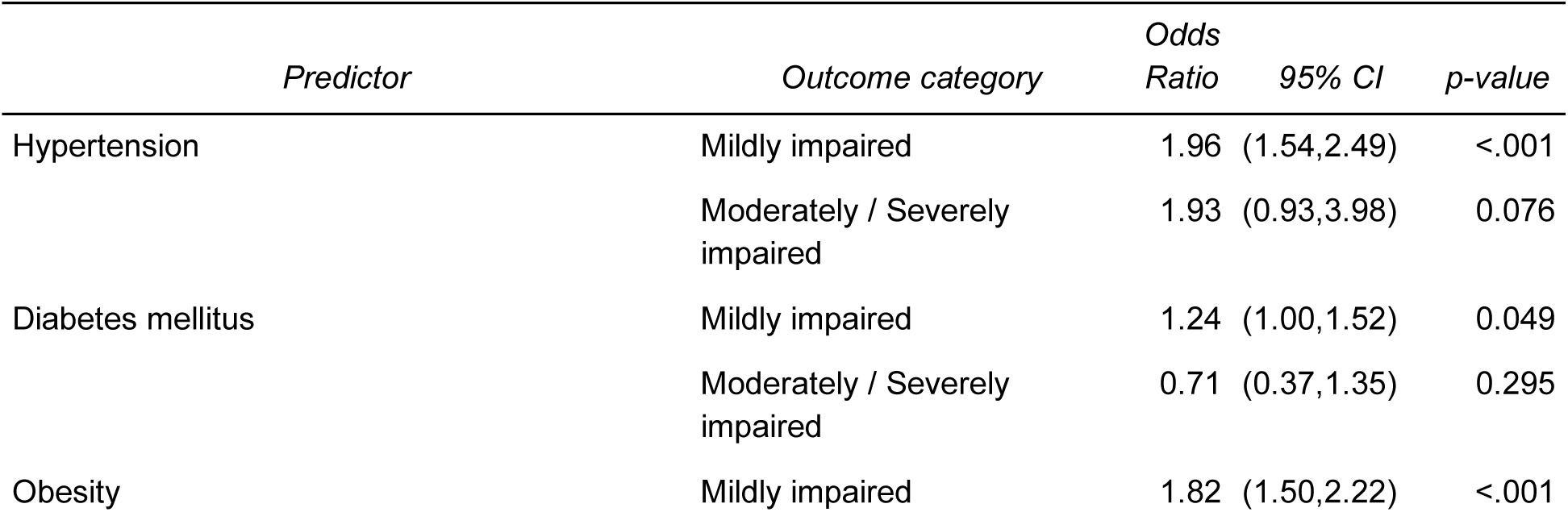
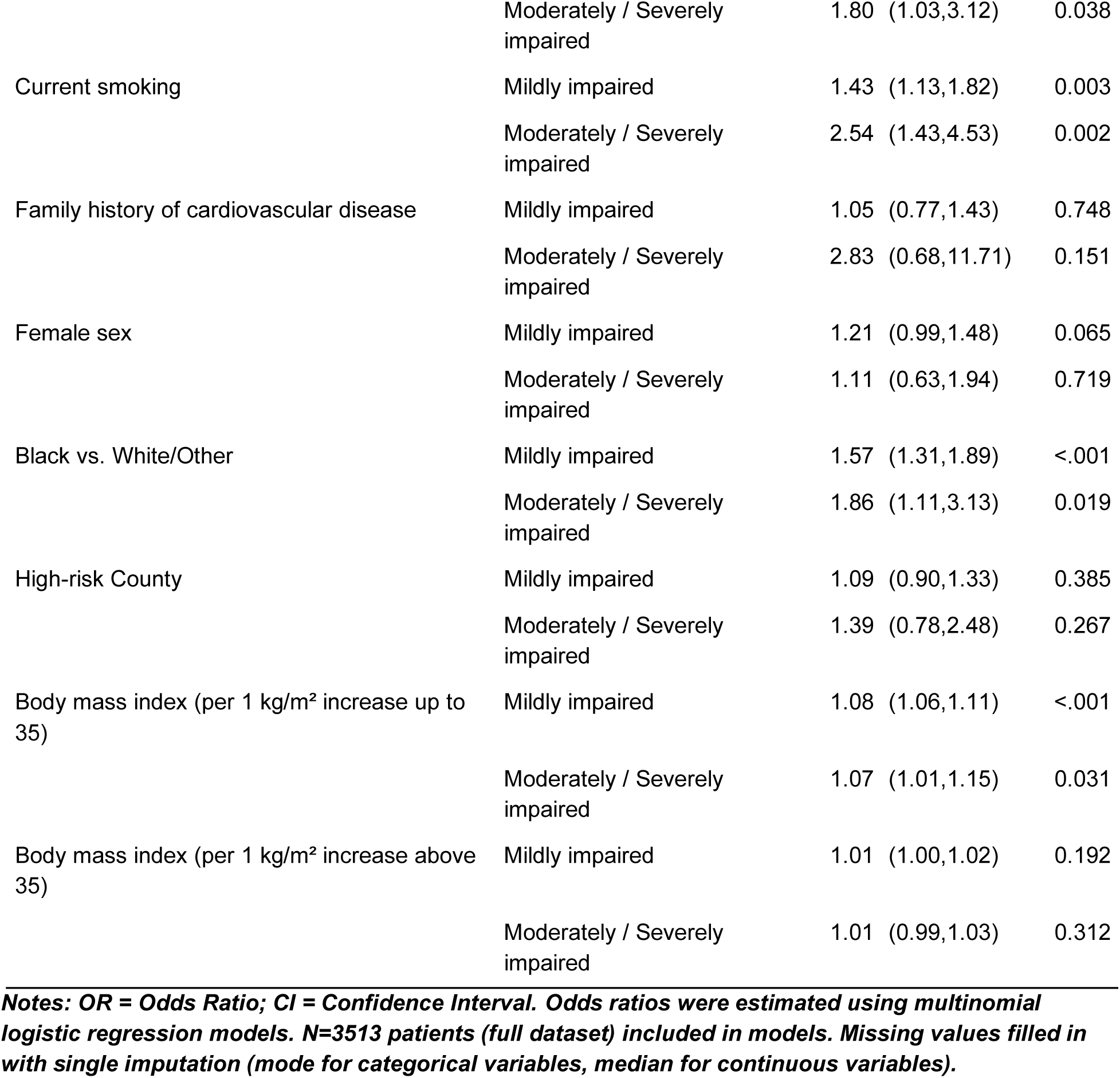
Multinomial Logistic Regression Results (Reference = Preserved, adjusted for age) For BSA-indexed (Primary Endpoint 2)

**Table 6a3:**
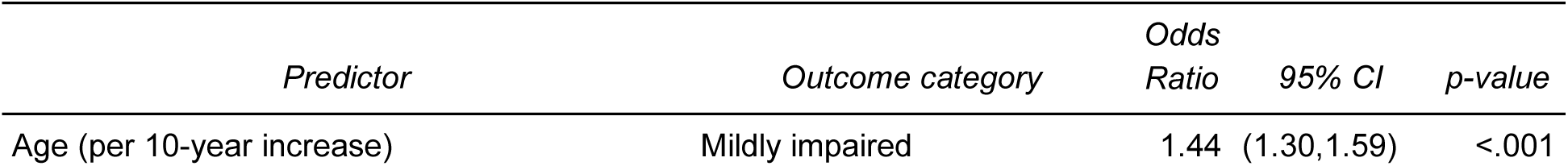
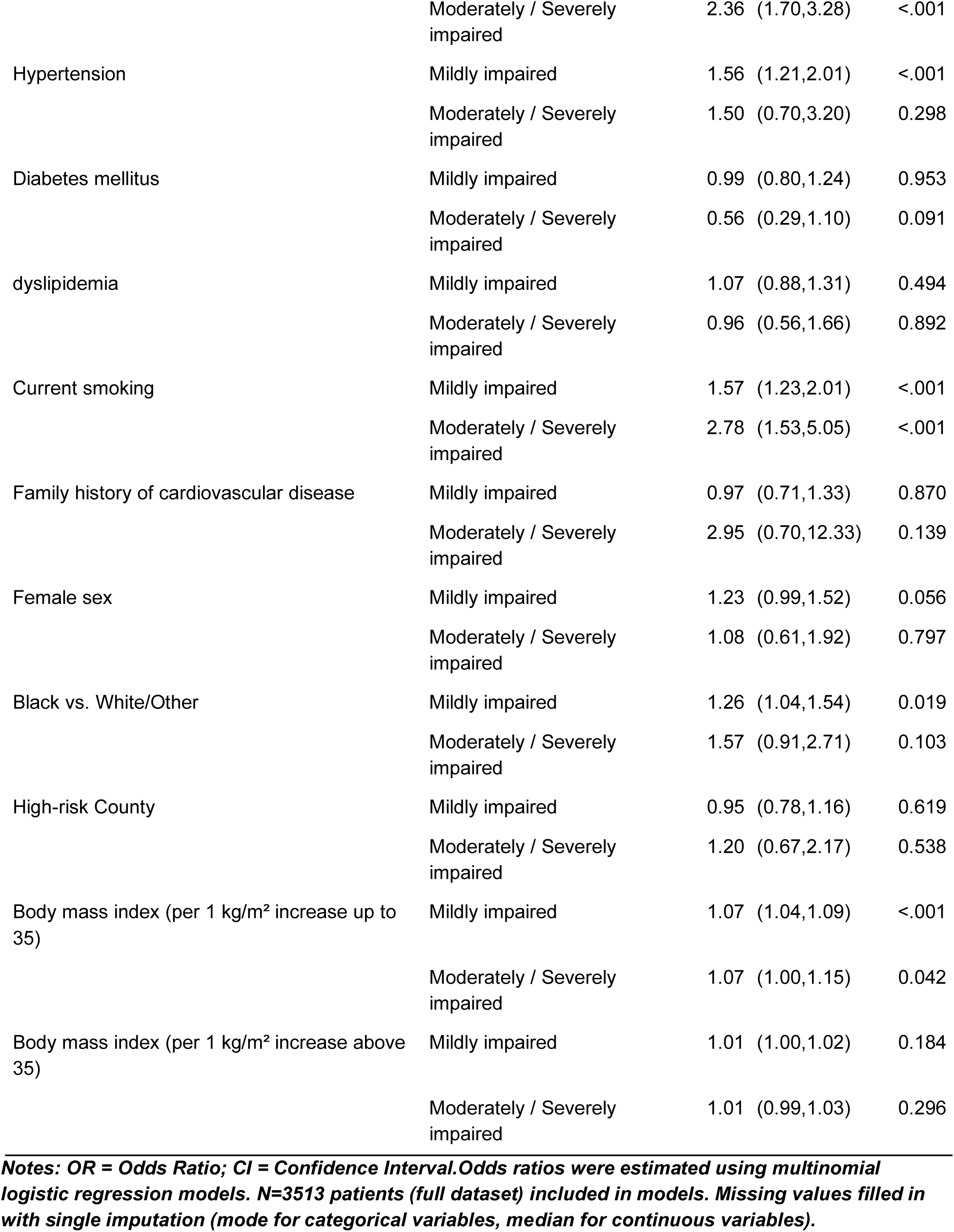
Multivariable multinomial Logistic Regression Results (Reference = Preserved, with BMI) For BSA-indexed (Primary Endpoint 2)

**Table 6b1:**
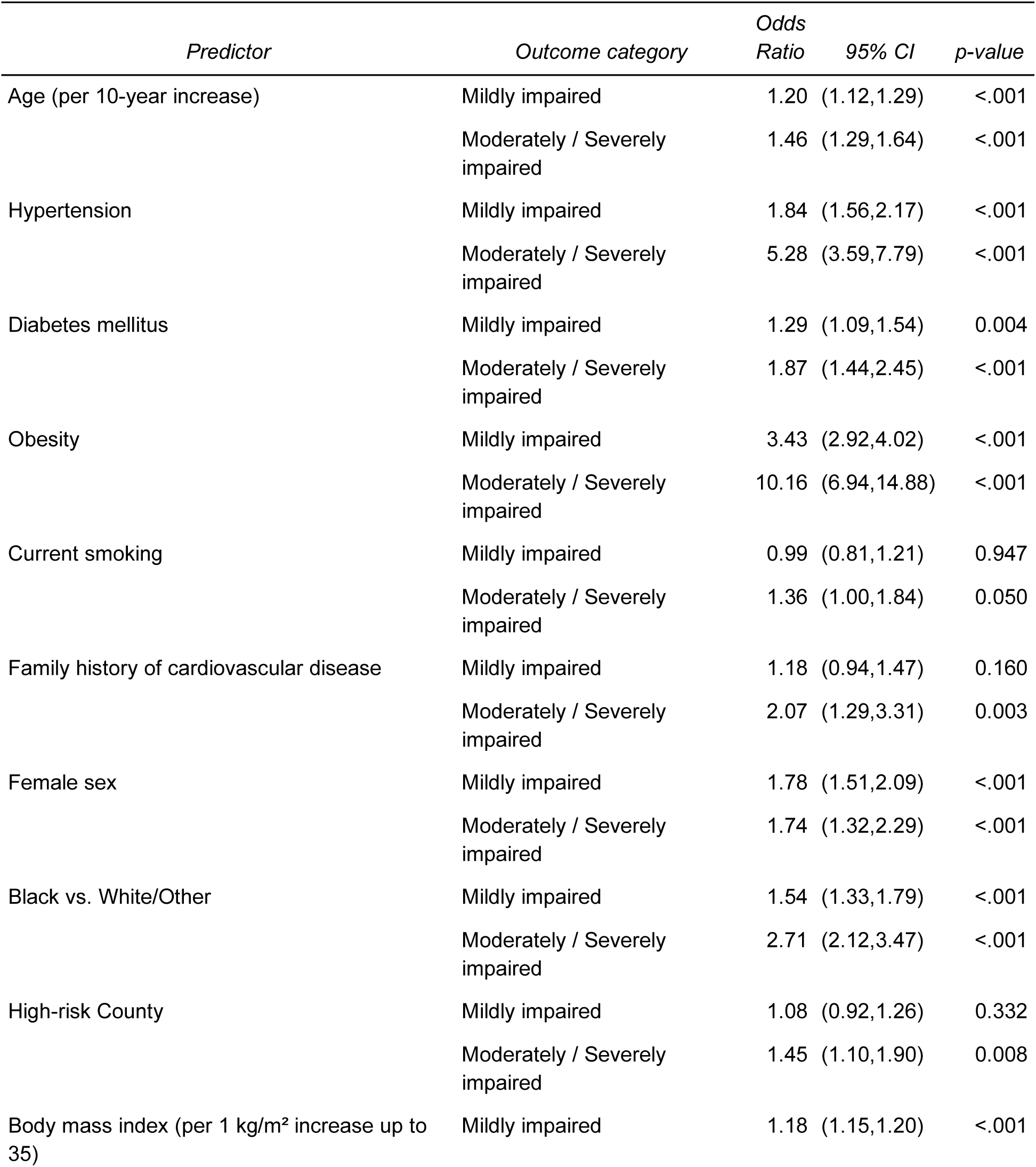
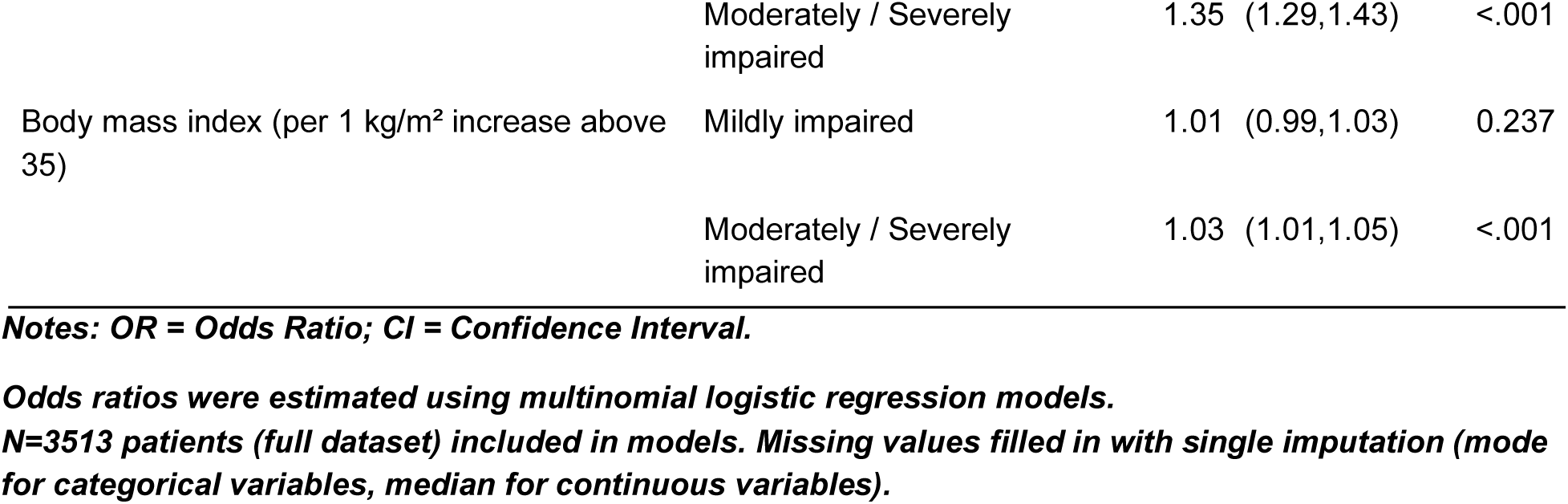
Univariable Multinomial Logistic Regression Results (Reference = Preserved) For height-indexed (Primary Endpoint 3)

**Table 6b2:**
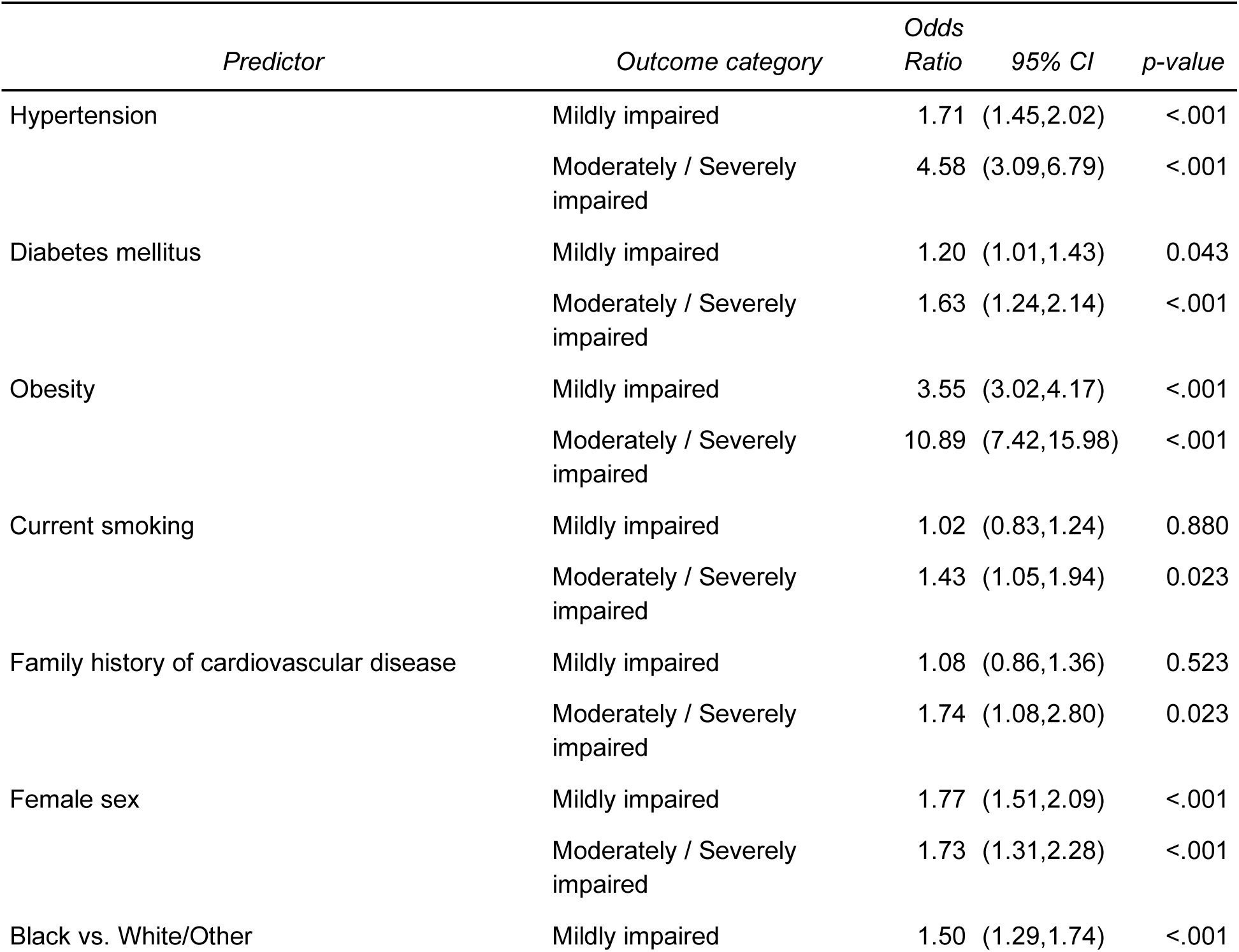
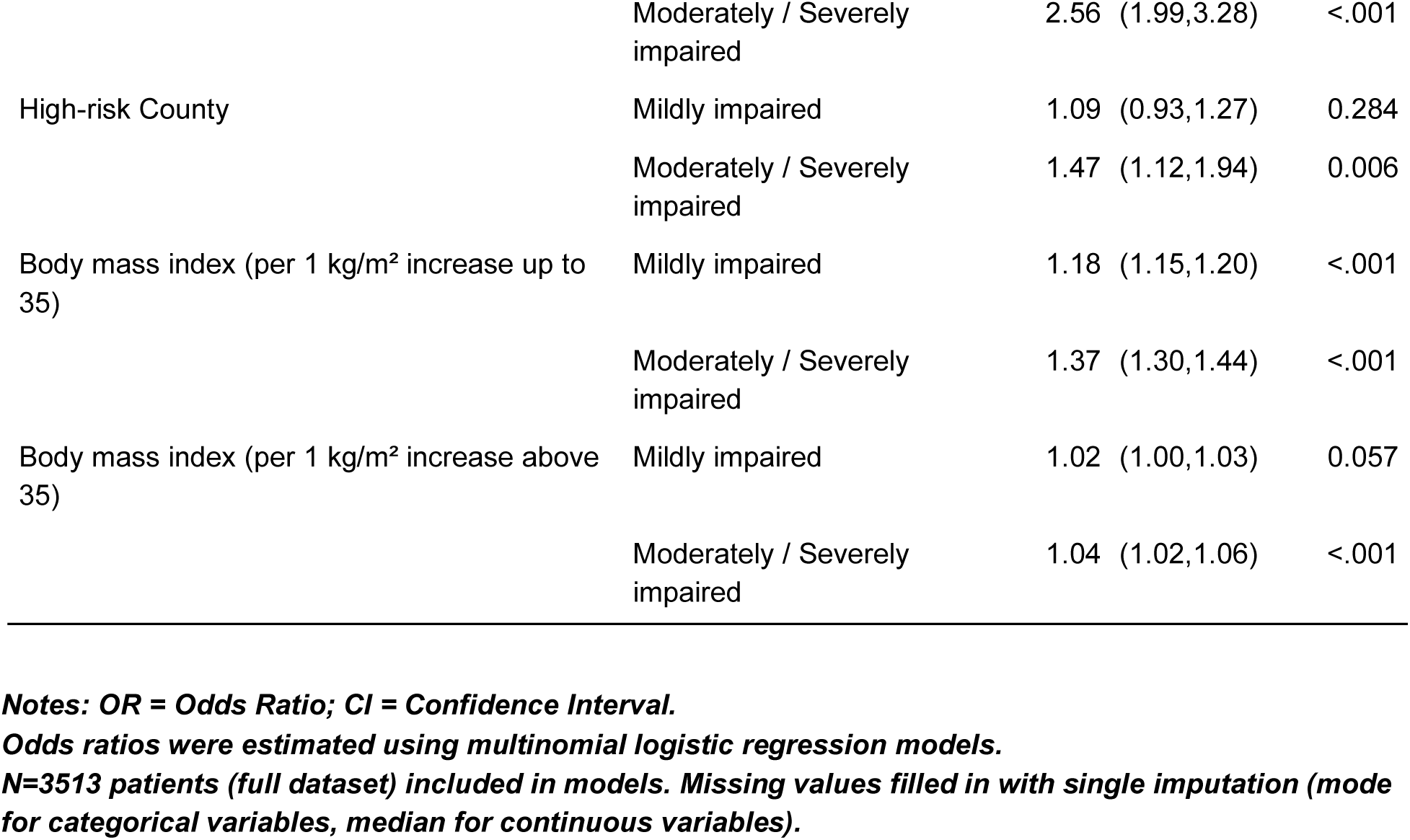
Multinomial Logistic Regression Results (Reference = Preserved, adjusted for age) For height-indexed (Primary Endpoint 3)

**Table 6b3:**
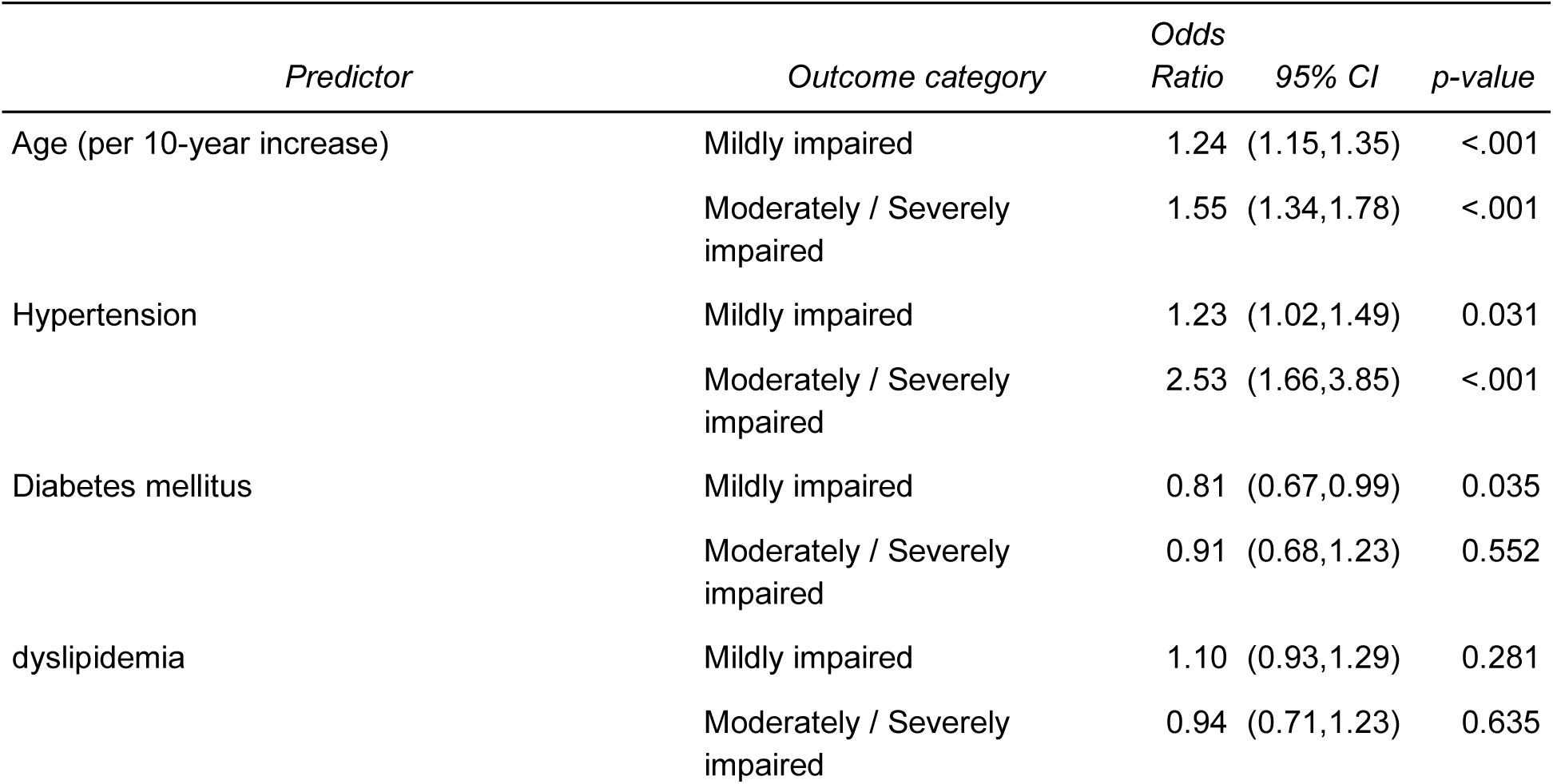
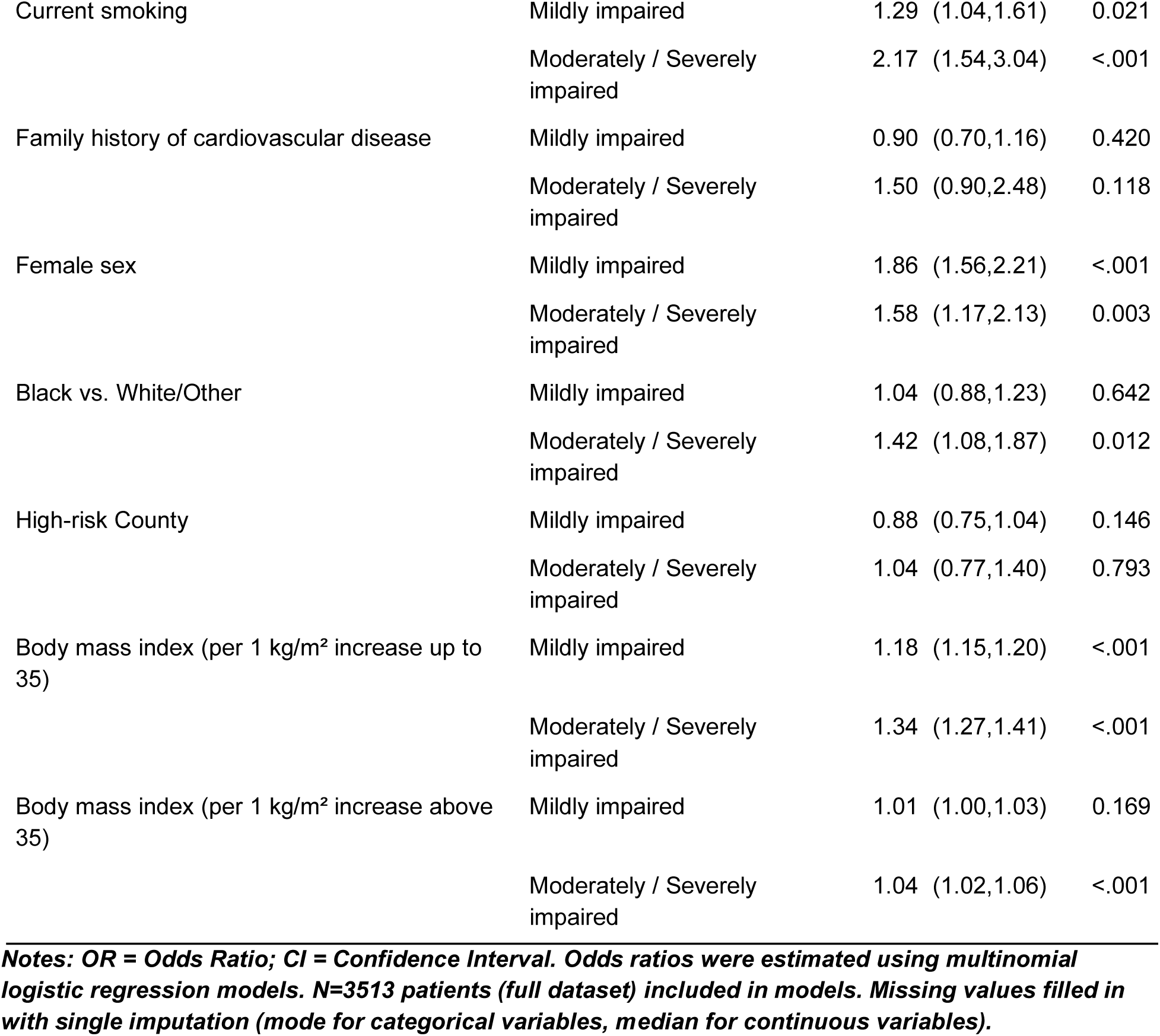
Multivariable multinomial Logistic Regression Results (Reference = Preserved, with BMI) For height-indexed (Primary Endpoint 3)

**Table 7:** Associations between elevated Biomarkers and LV-LA score adjusting for LVEF and PREVENT score

| Outcome | LA-LV Health Score | Comparison | Odds Ratio (95% CI) | P Value |
| --- | --- | --- | --- | --- |
| Troponin | LVM/BSA & LAV/BSA | Mildly impaired | 1.42 (0.94-2.15) | 0.094 |
|  |  | Moderately / Severely impaired | 1.33 (0.50-3.54) | 0.562 |
|  | LVM/H2.7 & LAV/H2.7 | Mildly impaired | 1.15 (0.82-1.60) | 0.413 |
|  |  | Moderately / Severely impaired | 1.30 (0.79-2.14) | 0.307 |
|  | *LVM/H2.7 & LAV/H2* | *Mildly impaired* | 1.56 (1.10-2.19) | 0.011 |
|  |  | *Moderately / Severely impaired* | 1.27 (0.73-2.22) | 0.394 |
|  | *LVM/H2.7 & LAV/H1.7* | *Mildly impaired* | 1.35 (0.97-1.88) | 0.073 |
|  |  | *Moderately / Severely impaired* | 1.30 (0.79-2.15) | 0.304 |
|  | *LVM/BSA & LAV/H2.7* | *Mildly impaired* | 0.94 (0.68-1.31) | 0.735 |
|  |  | *Moderately / Severely impaired* | 1.07 (0.55-2.08) | 0.842 |
|  | *LVM/BSA & LAV/H2* | *Mildly impaired* | 1.20 (0.85-1.70) | 0.307 |
|  |  | *Moderately / Severely impaired* | 1.03 (0.50-2.13) | 0.939 |
|  | *LVM/BSA & LAV/H1.7* | *Mildly impaired* | 1.06 (0.76-1.46) | 0.740 |
|  |  | *Moderately / Severely impaired* | 1.12 (0.58-2.18) | 0.727 |
| *NT proBNP* | *LVM/BSA & LAV/BSA* | *Mildly impaired* | 2.10 (1.19-3.70) | 0.011 |
|  |  | *Moderately / Severely impaired* | 1.14 (0.30-4.36) | 0.849 |
|  | *LVM/H2.7 & LAV/H2.7* | *Mildly impaired* | 1.18 (0.68-2.03) | 0.556 |
|  |  | *Moderately / Severely impaired* | 1.81 (0.91-3.59) | 0.092 |
|  | *LVM/H2.7 & LAV/H2* | *Mildly impaired* | 1.39 (0.81-2.41) | 0.234 |
|  |  | *Moderately / Severely impaired* | 2.01 (0.97-4.18) | 0.062 |
|  | *LVM/H2.7 & LAV/H1.7* | *Mildly impaired* | 1.16 (0.68-2.00) | 0.587 |
|  |  | *Moderately / Severely impaired* | 1.72 (0.86-3.45) | 0.128 |
|  | *LVM/BSA & LAV/H2.7* | *Mildly impaired* | 1.31 (0.77-2.20) | 0.316 |
|  |  | *Moderately / Severely impaired* | 2.31 (1.03-5.15) | 0.041 |
|  | *LVM/BSA & LAV/H2* | *Mildly impaired* | 1.71 (1.00-2.89) | 0.048 |
|  |  | *Moderately / Severely impaired* | 2.33 (0.97-5.57) | 0.058 |
|  | *LVM/BSA & LAV/H1.7* | *Mildly impaired* | 1.29 (0.77-2.17) | 0.339 |
|  |  | Moderately / Severely impaired | 2.16 (0.95-4.91) | 0.065 |

**Notes:** This analysis was run in the biomarker sub-study (N=970). Elevated Troponin defined as $\geq 6$ ng/L. Elevated NT proBNP defined as $\geq 125$ pg/mL. OR = Odds Ratio; CI = Confidence Interval. Odds ratios were estimated using logistic regression models (Reference = Preserved). PREVENT score had missing values (10%), single imputation was performed using median.

**Figure 1a:**
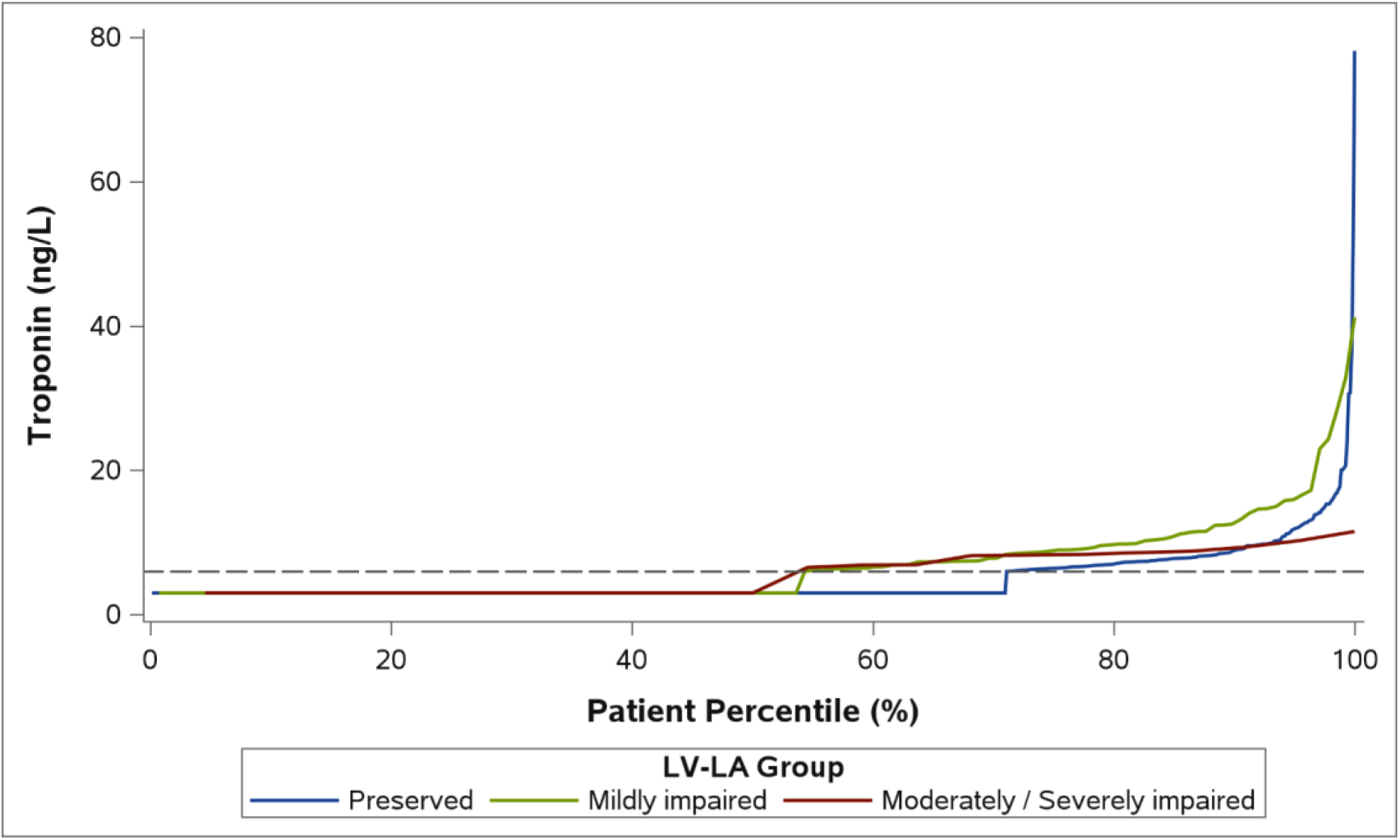
Cumulative Distribution of Troponin by LV-LA Group (BSA) Reference line indicates Undetectable Low < 6 ng/L

**Figure 1b:**
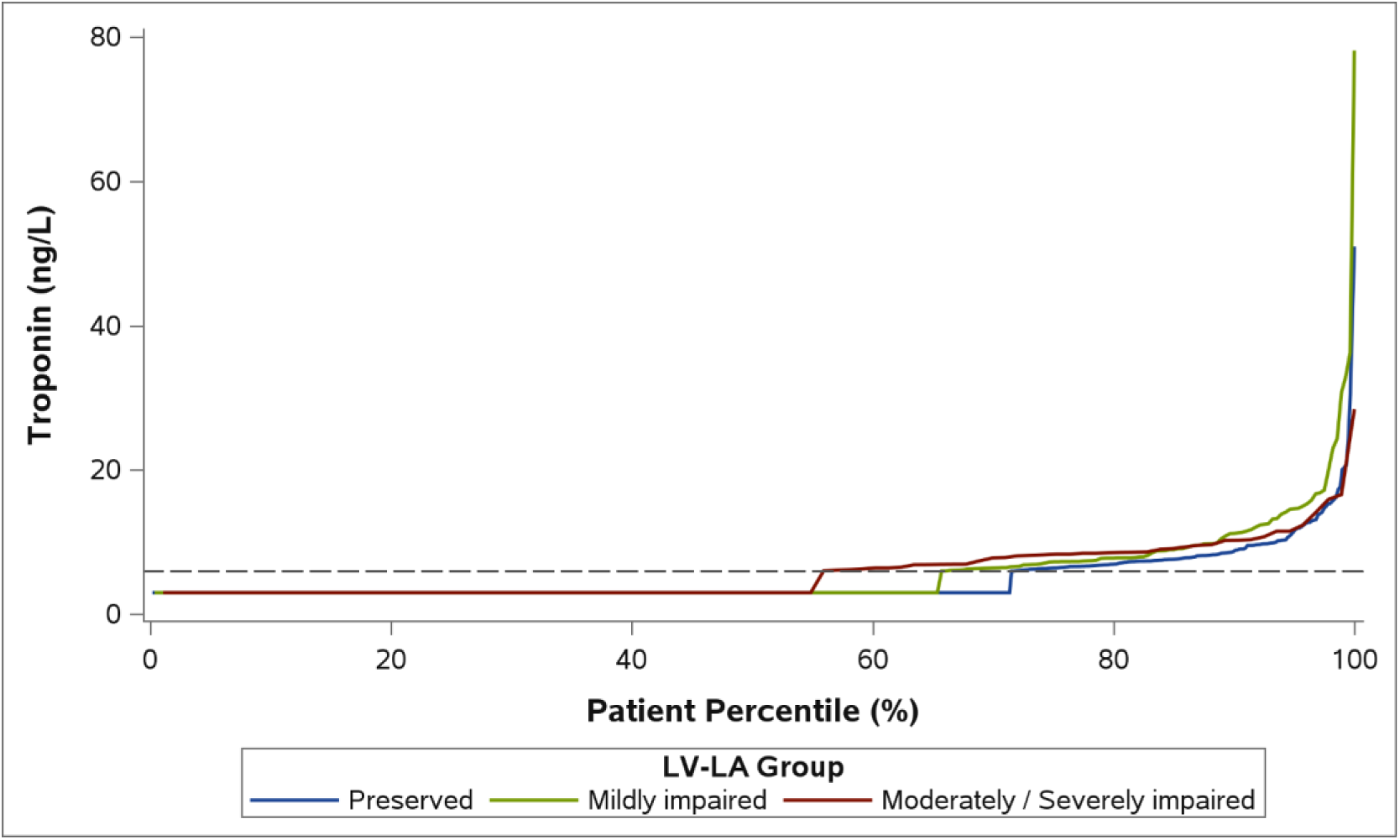
Cumulative Distribution of Troponin by LV-LA Group (LVM/H2.7 & LAV/H2.7) Reference line indicates Undetectable Low < 6 ng/L

**Figure 2a:**
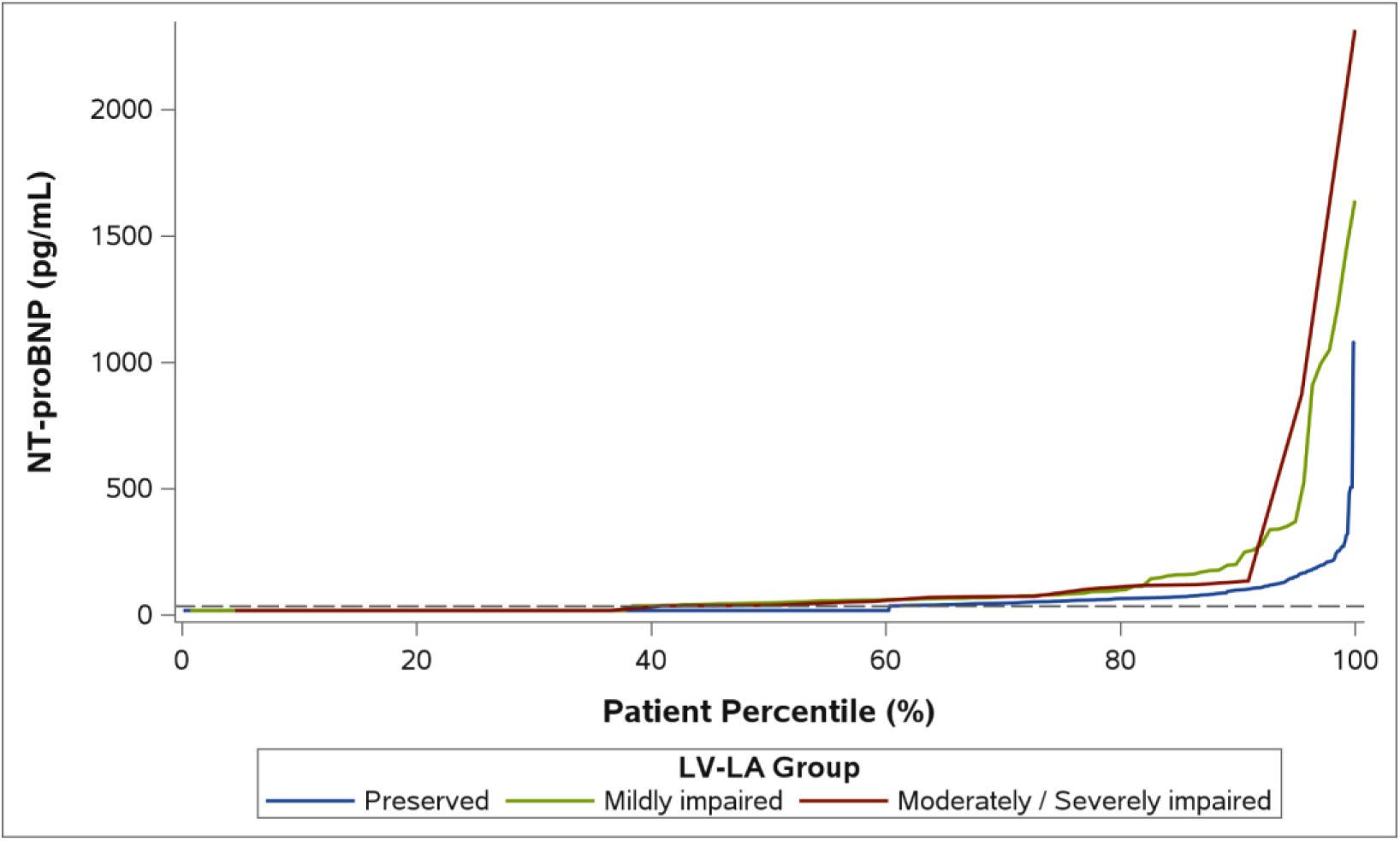
Cumulative Distribution of NT-proBNP by LV-LA Group (BSA) Reference line indicates Undetectable Low < 36 pg/mL

**Figure 2b:**
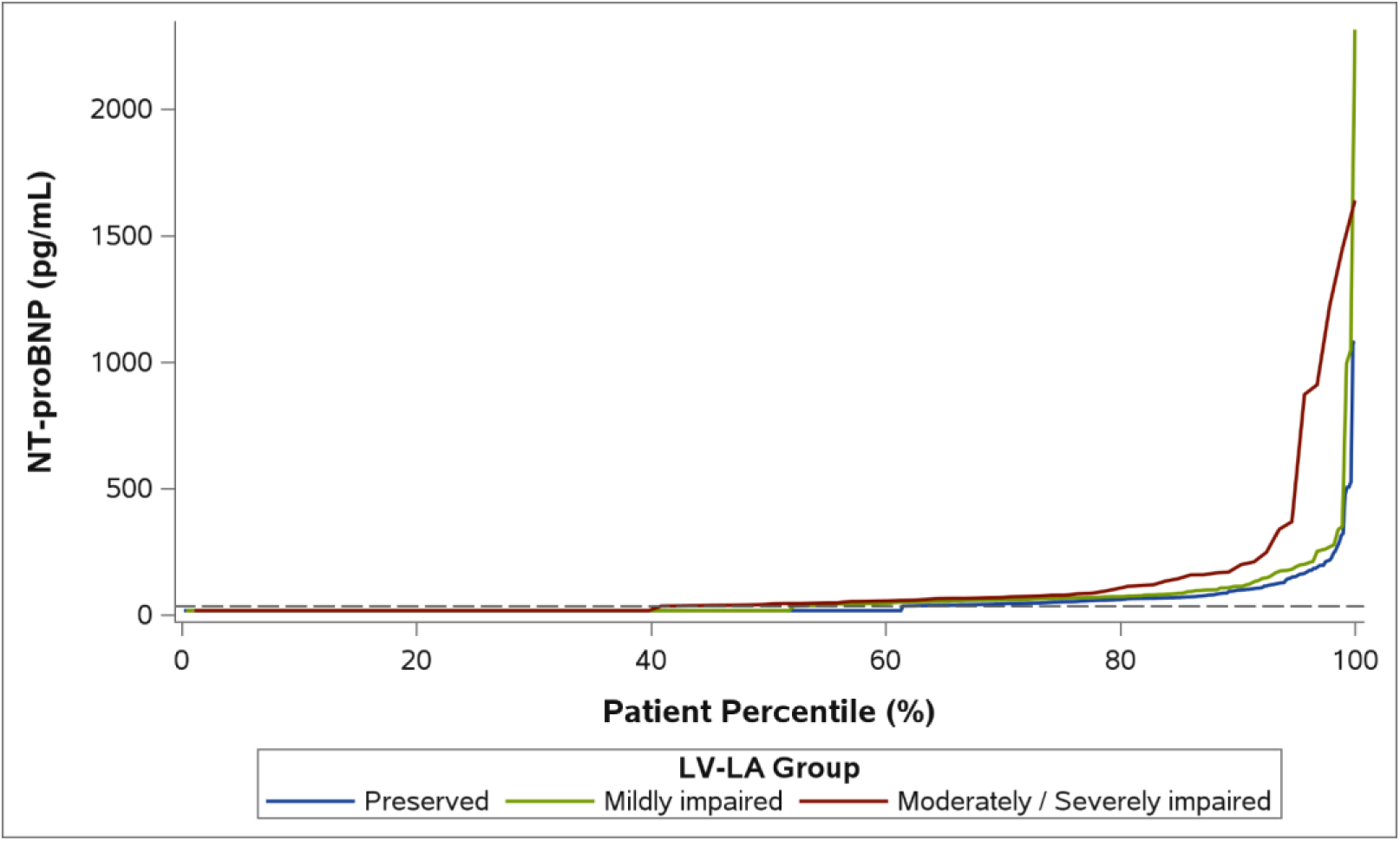
Cumulative Distribution of NT-proBNP by LV-LA Group (LVM/H2.7 & LAV/H2.7) Reference line indicates Undetectable Low < 36 pg/mL

**Figure 3:**
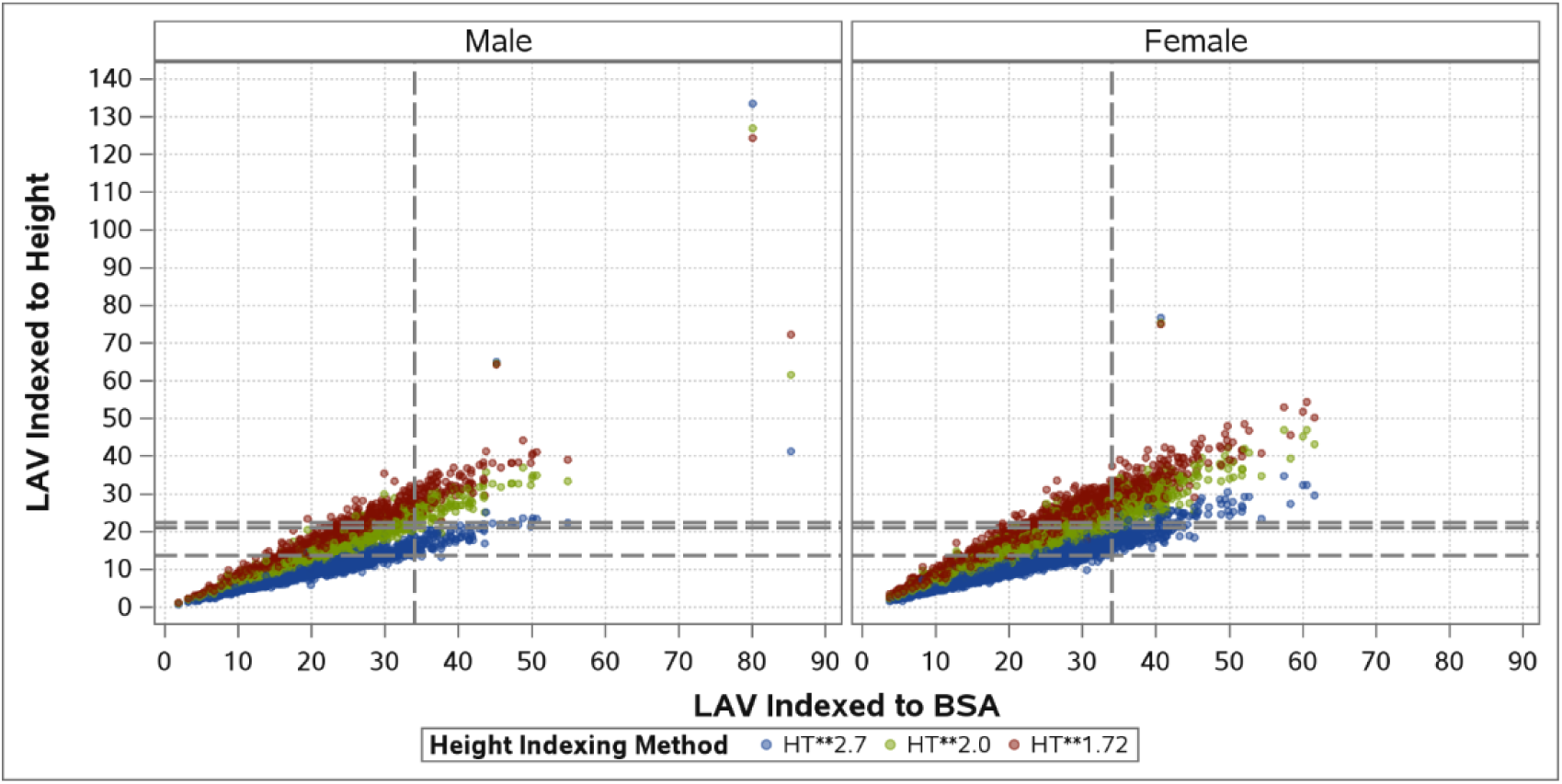
Comparison of LAV Indexing Methods stratified by Male and Female The horizontal reference lines indicate the normal cuts of 13.6, 21.0 and 22.3 for LAV Indexed to HT**2.7, HT**2.0 and HT**1.72 respectively The vertical reference line indicates the normal cut of 34 for LAV Indexed to BSA

## Data Availability

All data produced in the present study are available upon reasonable request to the authors

